# Temporal and Spatial Patterns of Snakebite Envenoming in Ghana, 2020-2025: A Nationwide Surveillance Analysis

**DOI:** 10.64898/2026.07.12.26357875

**Authors:** Eric Nyarko, Pascal Antwi, Ebenezer Boakye Amponsah, Lawrence Ofori-Boadu, Ebenezer Oduro-Mensah, Joseph Adjetey Oliver-Commey, Malik Haruna, Cynthia Serwaa, George Dadzie

## Abstract

Snakebite envenoming is a major neglected tropical disease disproportionately affecting rural populations in sub-Saharan Africa. In Ghana, evidence on the spatial and temporal distribution of risk remains limited, constraining targeted prevention and resource allocation. This study quantified district-level snakebite risk across Ghana, identified persistent hotspots and environmental drivers, and evaluated the relationship between snakebite burden and geographic access to treatment. Monthly district-level snakebite cases from Ghana’s District Health Information Management System (2020 to 2025) were analyzed across all 261 districts using a Bayesian spatio-temporal model incorporating spatial effects, a temporal random effect, and a space-time interaction, fitted via Integrated Nested Laplace Approximation. Environmental covariates including rainfall, temperature, humidity, and NDVI quantified associations with risk. Relative risks, exceedance probabilities, Local Indicators of Spatial Association, and geographic accessibility identified priority districts. Snakebite risk showed strong spatial clustering and temporal variation. Persistent high risk districts were concentrated in Upper West (Daffiama Bussie Issa, Wa East, Wa West, Sissala East), Savannah (Bole, Gonja), North East (Mamprugu Moagduri), Western North (Bia East), Bono (Banda), Oti (Krachi Nchumuru), Western (Wassa East), and Eastern Region (Nsawam Adoagyiri, Fanteakwa North), though patterns evolved. Fanteakwa North emerged as the highest risk district nationally in 2025. Humidity and temperature were associated with increased risk, while rainfall and NDVI showed no significant effect. High risk districts often had poor treatment access, revealing inequities. This first nationwide Bayesian spatio temporal assessment provides an evidence base for surveillance, antivenom distribution, and interventions supporting WHO’s 2030 snakebite reduction goals.

## 1 Introduction

Snakebite envenoming (SBE) is among the deadliest of the neglected tropical diseases (NTDs) with an estimated yearly 1.8 to 2.7 million envenomings and 81,000–138,000 deaths occur worldwide, with roughly 95% concentrated in low and middle-income countries (LMICs) [1], and recent Global Burden of Disease analyses identify it as a leading cause of mortality among the NTDs [2]. After a contested history on the global health agenda, it was reinstated as a category A NTD in 2017 [1], prompting the World Health Organization’s (WHO) 2019 roadmap target of halving snakebite death and disability by 2030—a target dependent on the spatially and temporally resolved surveillance that endemic regions conspicuously lack.

Sub-Saharan Africa (SSA) carries a disproportionate share of this burden. Annual incidence is estimated at 56 persons per 100,000 population from hospital data and up to 204 persons per 100,000 population from household surveys, with case-fatality between 2.8% and 11.6% [3], and West Africa alone accounts for approximately 320,000 disability-adjusted life years annually, around 91% of which derives from early mortality [4]. The principal culprit is Echis ocellatus, the West African carpet viper, whose haemotoxic venom causes severe hemorrhage and multiorgan failure [5], alongside the puff adder *(Bitis arietans)* and spitting cobras *(Naja spp.)*. This burden is compounded by chronic shortfalls in antivenom distribution, driven in part by poor epidemiological data on where and when need is greatest [5].

In Ghana, snakebite surveillance is based on health facility-reported data. Ghana’s District Health Information Management System (DHIMS-2) captures only aggregated counts without identifiers or species attribution [6], and existing national work has been descriptive, cataloging species and venom activity without quantitative spatial analysis [7]. Regional analyses exist—including a geospatial study of Savannah Region data for 2018–2022 [8]—but no nationwide Bayesian spatio-temporal risk model has been studied. These data pose a characteristic statistical challenge. District counts are modeled as Poisson variables whose mean is the product of an expected count and a relative risk (RR) [9], but crude rates are unstable at low counts and ignore spatial dependence. The modified Besag–York–Mollíe (BYM2) model decomposes log-risk into spatially structured and unstructured components [10], and its scaled reparameterization, BYM2, adds an interpretable mixing parameter and penalized-complexity priors, fitted via integrated nested Laplace approximation (INLA) [11, 12]. Purely spatial models, however, cannot capture temporal dynamics; Knorr-Held’s framework extends disease mapping to space and time through structured temporal trends and space–time interaction terms, allowing each district’s trajectory to differ [13], now standard in spatio-temporal mapping via INLA [14]. The resulting posteriors yield RRs with credible intervals (CrIs) and exceedance probabilities distinguishing confidently elevated districts from those merely appearing so, while global Moran’s I [15] and local indicators of spatial association (LISA) [16] formally test and localize hotspot clustering.

These models also accommodate covariates, explaining rather than merely mapping risk. A hierarchical Bayesian model in Nepal’s Terai identified vegetation (NDVI), distance to water, and poverty as principal predictors [9], and comparable geospatial approaches have delineated environmental risk hotspots in Iran [17] and Mexico [18]. A systematic review further confirms temperature, humidity, and rainfall as the climatic variables most consistently associated with snakebite incidence [19, 20]. These mechanisms are pertinent to Ghana, where rainfall, temperature, humidity, and vegetation jointly modulate snake activity and seasonal agricultural exposure [8, 20]. This study employed a Bayesian spatio-temporal BYM2 model with structured temporal and space–time interaction effects, applied to all 261 district-level health facility-reported snakebite data (2020–2025) from DHIMS-2, to estimate district RR with quantified uncertainty, identify statistically credible hotspots and their temporal evolution, healthcare accessibility, and quantify the environmental drivers of risk—rainfall, temperature, humidity and vegetation. This provides the first nationwide spatially and temporally explicit evidence base to guide targeted prevention and equitable antivenom distribution in Ghana.

## 2 Methods

### 2.1 Study Area

Ghana is a West African nation of approximately 238,535 km^2^ along the Gulf of Guinea, bordered by Ĉote d’Ivoire, Burkina Faso, and Togo. It is administratively divided into 16 regions, subdivided into 261 metropolitan, municipal, and district assemblies that constitute the units of analysis [21]. The country spans a pronounced north-south gradient, from the hot, semi-arid savanna of the north to the humid forests of the south, encompassing five agro-ecological zones. Rainfall is seasonal—bimodal in the south, unimodal in the north. These climatic and vegetative gradients structure both the distribution of medically important snakes [7], particularly the savanna-dwelling *Echis ocellatus* and *Bitis arietans*, and the agrarian livelihoods that bring rural populations into contact with them.

### 2.2 Data Source

District-level monthly snakebite case counts for 2020–2025 were obtained from the DHIMS-2, which records routinely reported cases as aggregated counts. Each record includes the health facility where the case was treated; because DHIMS-2 captures cases at the point of treatment rather than at the location of the bite or the patient’s residence, district counts reflect the treatment location. The names of facilities reporting snakebite cases were extracted from the surveillance records and geocoded to obtain their geographic coordinates, which were manually reviewed for positional accuracy; only facilities with verified coordinates were retained for the access analysis. District administrative boundaries (shapefile) were obtained from the Global Administrative Areas (GADM) database (https://gadm.org) [22] and projected to Universal Transverse Mercator (UTM) Zone 30N (EPSG:32630). District population estimates for 2020–2025 were derived from the WorldPop Constrained Individual Countries gridded dataset at 1 km resolution [23], extracted to district boundaries by summing pixel values. Monthly climatic covariates—maximum temperature ranges from (26.02 to 44.41) (*^◦^*C) and minimum temperature ranges (9.65 to 27.17) (*^◦^*C), rainfall (mm), and relative humidity (%)—were retrieved from the National Aeronautics and Space Administration POWER database [24] at district centroids via the nasapower R package. Monthly district-level NDVI was derived from the Moderate Resolution Imaging Spectroradiometer (MODIS) MOD13Q1 product (250 m, 16-day composites), with district-level mean values computed in Google Earth Engine [25] and the standard scale factor applied to yield values on the [−1, 1] range. Missing values due to cloud contamination were imputed using linear interpolation within each district’s time series.

### 2.3 Statistical Analysis

All analyses were performed in R version 4.5 [26], using sf and terra for spatial operations, spdep for neighborhood construction, and R-INLA for Bayesian inference [27]. Case records, population rasters, environmental layers, and district boundaries were imported and joined on a common identifier. District names were harmonized to a single canonical set through string standardization and a manually verified crosswalk, with metropolitan sub-districts dissolved to their parent assemblies. Population rasters were aggregated to districts by summing pixel values within each polygon, and environmental rasters were summarized to district-month means. Missing NDVI values due to cloud contamination in the MODIS composites were imputed using linear interpolation within each district series, and all covariates were standardized prior to modeling. Exploratory analyses summarized annual case totals and incidence rate per 100,000 population, and visualized regional variation using boxplots of district incidence. Spatial dependence was assessed using global Moran’s *I* per year, and a Kruskal-Wallis test was used to evaluate heterogeneity in risk across districts. Snakebite counts were modeled as Poisson-distributed with population as an offset, using a Bayesian spatio-temporal BYM2 model with a first-order random walk temporal trend and space-time interaction [11, 13]; models were compared using Deviance Information Criterion (DIC) and Watanabe-Akaike Information Criterion (WAIC). RR, 95% CrI, and exceedance probabilities, *P* (RR *>* 1), were derived from the posterior. Spatial clustering was assessed using local indicators of spatial association [16]. Distance to the nearest treatment facility was related to incidence using Spearman correlation.

### 2.4 Model Description

Monthly snakebite counts were modeled within a Bayesian hierarchical spatio-temporal framework. Let cases*_it_* denote the observed cases in district *i* (*i* = 1*, . . . ,* 261) and month *t* (*t* = 1*, . . . ,* 72; January 2020–December 2025). Counts followed a Poisson distribution as:

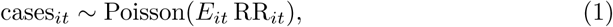

where RR*_it_* is the relative risk and *E_it_* the expected count obtained by internal standardization, *E_it_* = *n_it_ r*, with *n_it_* the monthly population exposure (annual district population divided by twelve) and *r* the overall study-period rate. The log RR was modeled as:

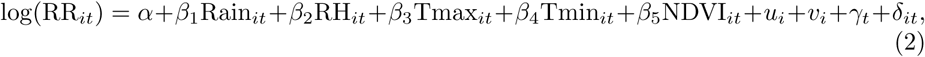

where *α* is the intercept and *β*_1_–*β*_5_ the coefficients of the standardized environmental covariates: rainfall, relative humidity (RH), maximum temperature (Tmax), minimum temperature (Tmin), and NDVI. These covariates are interpretable as the change in log-risk per one standard-deviation increase. The district effects *u_i_* and *v_i_* were specified jointly using the BYM2 reparameterization [11], which combines a structured intrinsic conditional autoregressive component [10] and an unstructured component through a single precision and a mixing parameter *ϕ* ∈ [0, 1], the proportion of district-level variance that is spatially structured. Adjacency was defined by the first-order queen contiguity, with isolated districts joined to their nearest neighbor. The temporal trend (*γ_t_*) was a first-order random walk, and the space-time interaction (*δ_it_*) followed the Type I specification of Knorr-Held [13].

All parameters received weakly informative priors. Penalized-complexity priors [12] were placed on the BYM2, temporal, and interaction precisions (P(*σ >* 1) = 0.01) and on the mixing parameter (P(*ϕ <* 0.5) = 0.5); the intercept and coefficients received vague Gaussian priors. Models were fitted by integrated nested Laplace approximation (INLA) [27]. Structures of increasing complexity (spatial; +temporal; +covariates; +interaction) and Poisson versus negative binomial likelihoods were compared by DIC and WAIC. The Poisson model was retained for inference, as it achieved the lowest DIC and WAIC; the negative binomial specification yielded comparable fit, and the two produced consistent covariate and risk estimates. RRs were summarized as posterior means with 95% CrIs, and exceedance probabilities P(RR*_it_ >* 1) identified districts with credibly elevated risk.

### 2.5 Risk mapping and cluster detection

To visualize the observed burden, crude district incidence was computed as the total reported cases divided by the district population and expressed per 100,000 person-years as:

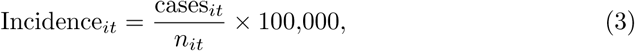

and mapped by year. From the fitted spatio-temporal model, posterior district-level RRs were summarized as posterior means as:

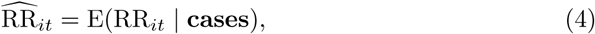

and mapped on a diverging scale centered on unity, where RR = 1 denotes risk equal to the national average; values above 1 indicate elevated risk, and values below 1 indicate lower-than-average risk. Because a high posterior mean alone does not establish that risk is genuinely elevated, exceedance probabilities were computed from the posterior marginals to quantify the certainty of elevated risk as:

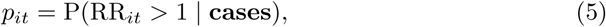

and districts with *p_it_ >* 0.95 were classified as credible high-risk areas [11]. This probabilistic criterion distinguishes districts whose elevated risk is supported by strong posterior evidence from those that appear elevated owing to sampling variability.

Local spatial clustering was assessed using Local LISA based on the local Moran’s *I* statistic [16] as:

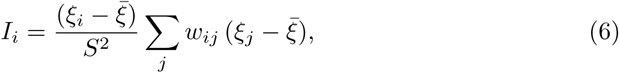

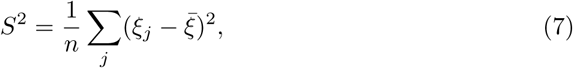

where *ξ_i_* is the relative risk in district *i*, *ξ* is the mean across districts, *w_ij_* the row-standardized first-order queen-contiguity spatial weight between districts *i* and *j*, and *n* the number of districts. Statistical significance was evaluated at the 0.05 level, and each district was assigned to one of five categories: high values surrounded by high values (high-high) and low surrounded by low (low-low), representing spatial clusters; high-low and low-high, representing spatial outliers; and non-significant. The local statistic was computed for each year to examine the temporal stability of identified clusters.

### 2.6 Geographic access to treatment

Geographic access to snakebite treatment was assessed using the geocoded locations of health facilities that reported snakebite cases. Facility names were geocoded and manually verified for positional accuracy, and only facilities with confirmed coordinates were retained. Facility locations and district boundaries were projected to the UTM zone 30N coordinate system (EPSG:32630), and the Euclidean distance from a representative interior point of each district to its nearest treatment facility was computed. The association between district snakebite incidence and distance to the nearest facility was evaluated using the Spearman rank correlation coefficient, which is robust to the skewed distribution of incidence. Districts combining above-median incidence with above-median distance to care were identified as priority underserved areas.

### 2.7 Ethical Consideration

As part of the Integrated Disease Surveillance and Response, snakebite cases are routinely reported in DHIMS-2. Consequently, the data used in this study were secondary data. We confirm that no personally identifiable or confidential information was accessed or used at any stage of the analysis.

## 3 Results

The results are presented in four parts: the overall burden and exploratory spatial tests; the spatio-temporal model fit and environmental drivers; the mapping of risk, hotspots, and their temporal evolution, and the geographic accessibility of treatment. Together, they trace a consistent geography of risk and its alignment with gaps in access to care.

### 3.1 Descriptive Statistics and Spatio-temporal Test

#### 3.1.1 Descriptive Statistics

Table 1 presents annual snakebite cases and incidence rates in Ghana from 2020 to 2025. Total cases declined from 11,084 in 2020 to 9,586 in 2025, while incidence per 100,000 decreased correspondingly from 2.94 to 2.31. Both cases and incidence declined fairly consistently over the period, with a slight increase in 2023 (10,545 cases; 2.64 per 100,000) relative to 2022 (10,054 cases; 2.56 per 100,000), then continued to decline through 2024 (10,463 cases; 2.59 per 100,000) and into 2025.

**Table 1.** Annual Snakebite Cases and Incidence Rates in Ghana, 2020–2025.

| Year | Total Cases | Incidence per 100,000 |
| --- | --- | --- |
| 2020 | 11,084 | 2.94 |
| 2021 | 10,783 | 2.80 |
| 2022 | 10,054 | 2.56 |
| 2023 | 10,545 | 2.64 |
| 2024 | 10,463 | 2.59 |
| 2025 | 9,586 | 2.31 |

#### 3.1.2 Spatio-temporal Test

We assessed whether snakebite risk varied across districts and whether it was spatially structured. Table 2 presents spatial heterogeneity and dependence tests. The Kruskal-Wallis statistic was 8530.8 (*p <* 0.001), indicating significant heterogeneity in district-level snakebite risk. The Global Moran’s I was 0.4817 (*p* = 0.001), indicating significant positive spatial autocorrelation, confirming that snakebite incidence is non-randomly distributed and clusters spatially across Ghana’s districts.

**Table 2.** Spatial Heterogeneity and Dependence Across Districts.

| Measure | Statistic | p-value |
| --- | --- | --- |
| Kruskal-Wallis Test | 8530.8 | < 0.001 |
| Global Moran's I | 0.4817 | 0.001 |

The annual Global Moran’s I statistics for snakebite incidence across districts are presented in Table 3. Spatial autocorrelation was significant in every year (*p* = 0.001), with Moran’s I ranging from 0.301 in 2020 to a peak of 0.469 in 2023. Values were 0.418 in 2021, 0.400 in 2022, declining to 0.372 in 2024 before rising slightly to 0.386 in 2025. This indicates persistent, though fluctuating, spatial clustering of snakebite risk across districts from 2020 to 2025.

**Table 3.** Moran’s I Statistics by Year for Snakebite Incidence Across Districts.

| Year | Moran's I | p-value |
| --- | --- | --- |
| 2020 | 0.301 | 0.001 |
| 2021 | 0.418 | 0.001 |
| 2022 | 0.400 | 0.001 |
| 2023 | 0.469 | 0.001 |
| 2024 | 0.372 | 0.001 |
| 2025 | 0.386 | 0.001 |

##Fig. 1 shows district-level snakebite incidence by region, ordered by descending median. The Savannah region exhibited the highest median incidence (∼700 persons per 100,000 population), followed by the Upper West region (∼550 persons per 100,000 population) with the widest interquartile ranges, indicating substantial within-region district variation. Western North, Upper East, and North East showed moderate, more dispersed incidence. Southern regions—Ashanti, Central, Volta, and Greater Accra—had the lowest medians and narrowest ranges. Several regions exhibited high-incidence outliers, notably the Northeast, Bono, and Eastern, reflecting localized district-level extremes.

**Fig. 1.**
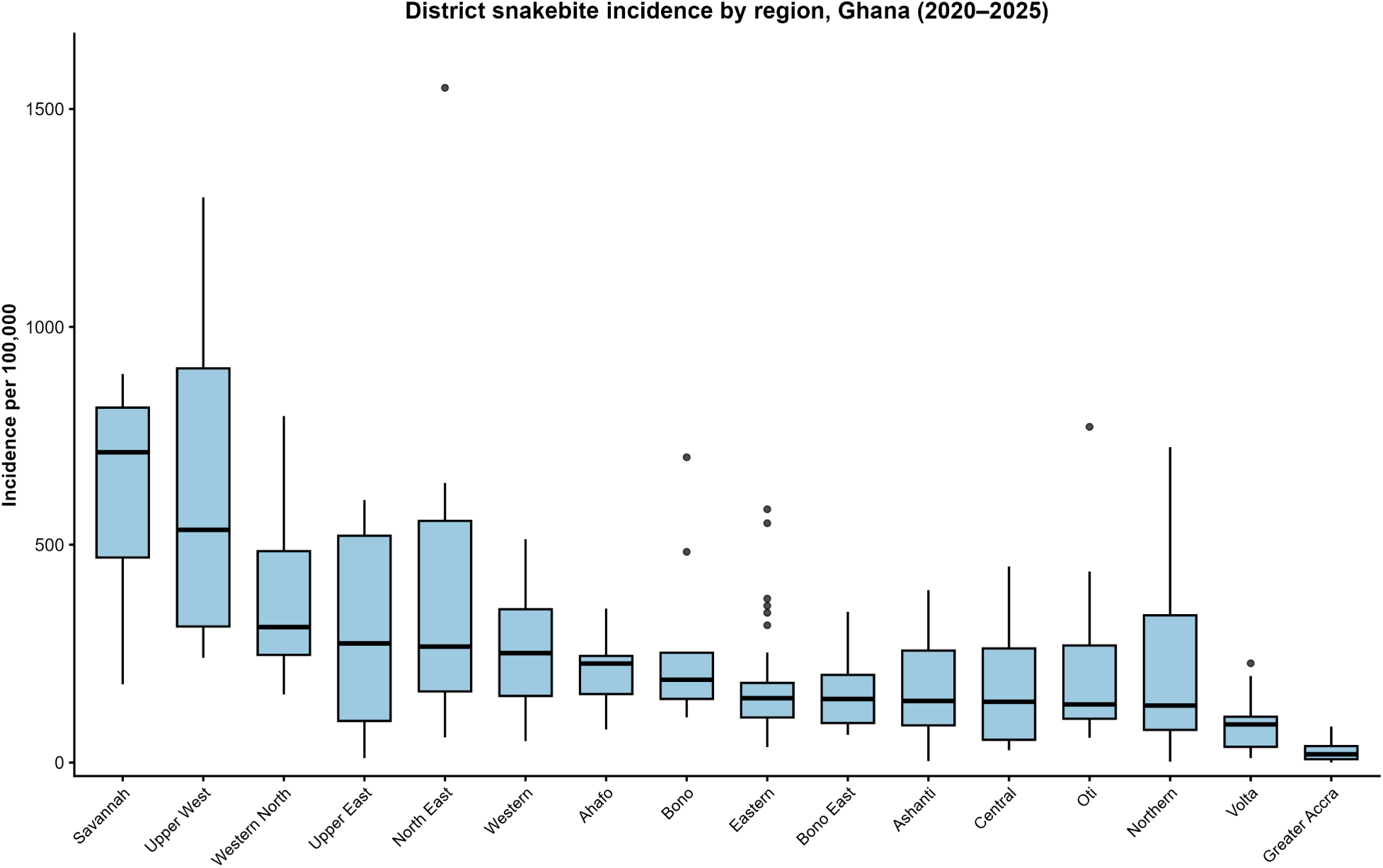
Distribution of district-level snakebite incidence (cases per 100,000 population) by region, Ghana, 2020–2025. Boxes show the median and interquartile range (IQR) of district-level incidence values within each region; whiskers extend to 1.5 *×* IQR, and points beyond this range are plotted as outliers. Regions are ordered by descending median incidence. Population denominators were derived from gridded population estimates aggregated to the district level.

### 3.2 Spatio-Temporal Model Fit

We compared models of increasing complexity, and alternative likelihoods, to identify the specification best supported by the data.

#### 3.2.1 Spatio-Temporal Model Selection

Table 4 compares likelihood specifications for the full spatio-temporal model. The Poisson model yielded DIC = 68019 and WAIC = 67863, while the negative binomial model yielded DIC = 68258 and WAIC = 67963. The Poisson likelihood thus showed marginally better fit (ΔDIC = 239, ΔWAIC = 100) and was retained for inference.

**Table 4.** Comparison of likelihood specifications for the full spatio-temporal model.

| Likelihood | DIC | WAIC |
| --- | --- | --- |
| Poisson | 68019 | 67863 |
| Negative Binomial | 68258 | 67963 |

Table 5 compares Poisson spatio-temporal models of increasing complexity using DIC, WAIC, and effective number of parameters (*p_D_*). The spatial-only model (*M*_0_) yielded DIC = 81005, WAIC = 99952, *p_D_* = −4479. Adding a temporal trend (*M*_1_) reduced fit indices to DIC = 76362, WAIC = 99140, *p_D_* = −4414. Incorporating covariates (*M*_2_) further improved fit (DIC = 76153, WAIC = 97739, *p_D_* = −4276). The largest improvement occurred with the space–time interaction term (*M*_3_), yielding DIC = 68030 and WAIC = 67878, alongside a markedly higher *p_D_* = 8948, reflecting substantially increased model complexity and the best overall fit among the four specifications.

**Table 5.** Comparison of Poisson spatio-temporal models of increasing complexity.

| Model | DIC | WAIC | $p_D$ |
| --- | --- | --- | --- |
| M0: Spatial only | 81005 | 99952 | -4479 |
| M1: + Temporal | 76362 | 99140 | -4414 |
| M2: + Covariates | 76153 | 97739 | -4276 |
| M3: + Space-time interaction | 68030 | 67878 | 8948 |

#### 3.2.2 Fixed and Random Effects Estimates

We examined the environmental drivers of risk and the decomposition of residual variation into its spatial, temporal, and interaction components. Table 6 presents incidence rate ratios (IRR) with 95% CrI for environmental covariates from the Bayesian spatio-temporal model. Relative humidity (IRR = 1.131, 95% CrI: 1.073–1.192), maximum temperature (IRR = 1.110, 95% CrI: 1.050–1.172), and minimum temperature (IRR = 1.119, 95% CrI: 1.071–1.168) were significantly associated with increased snakebite risk, as their CrI excluded 1. Rainfall (IRR = 0.997, 95% CrI: 0.972–1.023) and NDVI (IRR = 1.027, 95% CrI: 0.999–1.057) were not statistically significant, with CrI spanning unity. The intercept (IRR = 0.524, 95% CrI: 0.495–0.554) reflects baseline risk at standardized covariate values of zero.

**Table 6.** Incidence rate ratios for environmental covariates from the Bayesian spatio-temporal model.

| Variable | IRR (95% CrI) |
| --- | --- |
| Intercept | 0.524 (0.495, 0.554) |
| Rainfall | 0.997 (0.972, 1.023) |
| Relative humidity | 1.131 (1.073, 1.192) |
| Maximum temperature (26.02 to 44.41°C) | 1.110 (1.050, 1.172) |
| Minimum temperature (9.65 to 27.17°C) | 1.119 (1.071, 1.168) |
| Normalized Difference Vegetation Index | 1.027 (0.999, 1.057) |

Table 7 presents posterior estimates of the random-effect hyperparameters from the Bayesian spatio-temporal model. The spatial precision (BYM2) was estimated at 0.811 (95% CrI: 0.627–1.036), with the mixing parameter *ϕ* = 0.840 (95% CrI: 0.625–0.954), indicating that the majority of district-level variance was spatially structured rather than unstructured. The temporal precision (RW1) was substantially higher at 16.010 (95% CrI: 10.477–23.412), reflecting a smoother temporal trend, while the space–time interaction precision was 2.274 (95% CrI: 2.176–2.375), indicating moderate variability in district-specific temporal trajectories.

**Table 7.** Posterior estimates of the random-effect hyperparameters from the Bayesian spatio-temporal model.

| Parameter | Estimate (95% CrI) |
| --- | --- |
| Precision (spatial, BYM2) | 0.811 (0.627, 1.036) |
| $\phi$ (spatial structured prop.) | 0.840 (0.625, 0.954) |
| Precision (temporal, RW1) | 16.010 (10.477, 23.412) |
| Precision (space–time interaction) | 2.274 (2.176, 2.375) |

### 3.3 Risk and Hotspot Mapping

We map the geography of snakebite risk through four complementary lenses: observed incidence, model-based RR, posterior exceedance probability, and local spatial clustering.

#### 3.3.1 Incidence Rate Mapping

Fig. 2 illustrates the observed annual snakebite incidence per 100,000 population across Ghana’s 261 districts for each year from 2020 to 2025 (see Table A1 in the Appendix). The purpose of this presentation is to inform public health researchers, epidemiologists, policymakers, and health officials about spatial and temporal patterns in snakebite incidence, to aid targeted intervention planning. The incidence rates are classified into five categories: 0-6.4, 6.4-16.7, 16.7-31.5, 31.5-57.2, and *>* 57.2 persons per 100,000 population. The resulting spatial distribution shows that each incidence category includes districts from various ecologically distinct regions across the country.

**Fig. 2.**
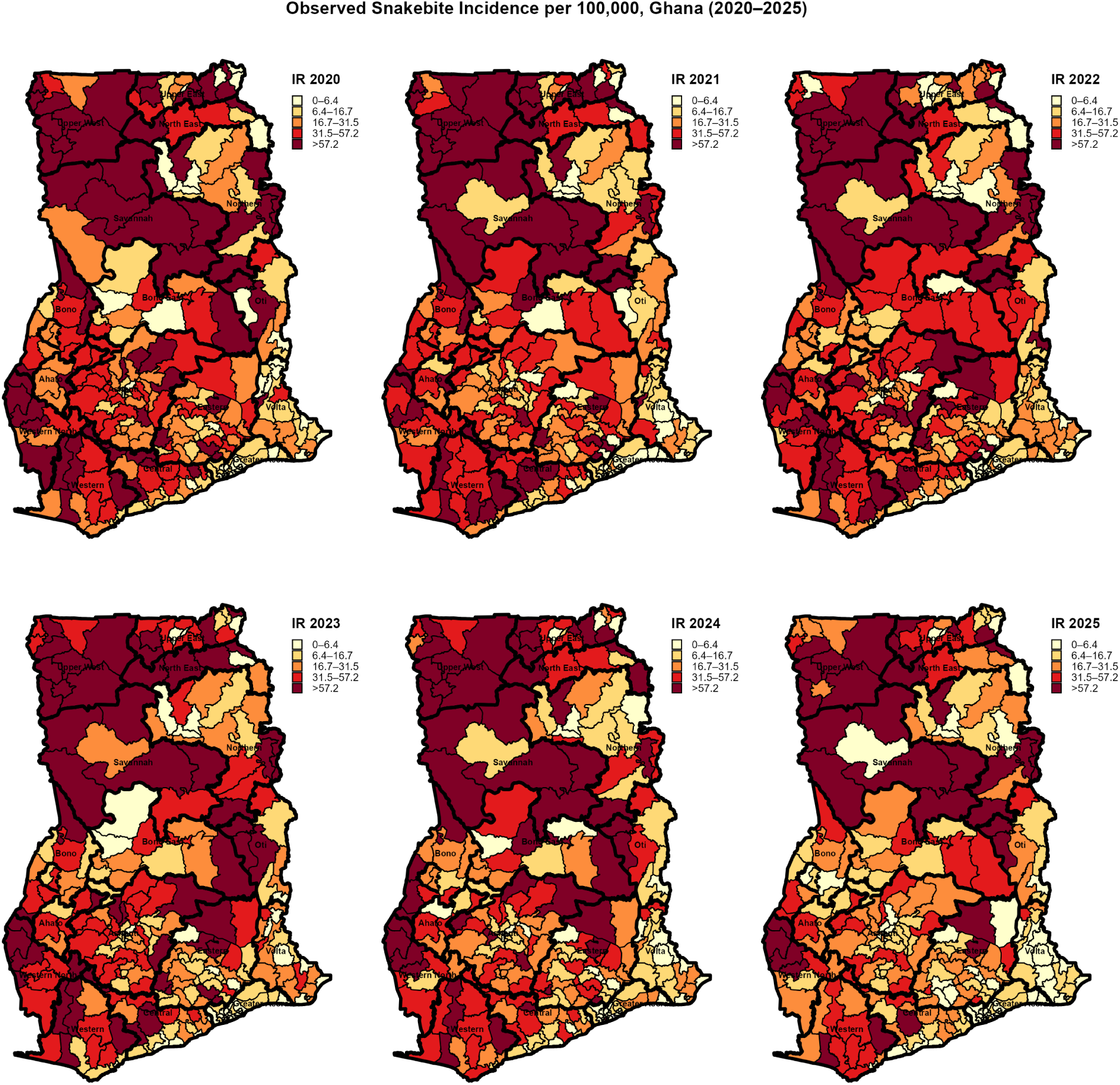
Observed snakebite incidence per 100,000 population across the 261 districts of Ghana, by year (2020–2025).

From 2020 to 2025, the Mamprugu Moagduri district in the North East Region consistently occupied the highest incidence tier, with rates of 122.4, 251.5, 424.7, 367.2, 215.9, and 165.6 persons per 100,000 population, peaking in 2022. Notably, this district’s persistently high incidence underscores its importance for targeted interventions. Daffiama Bussie Issa in the Upper West Region reported incidences ranging from 153.2 to 308.0, peaking in 2021. Wa East, also in the Upper West Region, had incidences from 69.8 to 219.5, peaking in 2022, while Wa West ranged from 109.2 to 201.3, peaking in 2021. In the Savannah Region, North East Gonja exhibited rates from 70.2 to 256.5, with the 2020 value being the highest district-level incidence recorded nationally that year, surpassing Bawku West in the Upper East Region, which reported 245.1. Banda in the Bono Region had rates varying from 43.3 to 183.0 from 2020 to 2025. Krachi Nchumuru in the Oti Region consistently reported rates above 57.2, ranging from 99.0 to 161.1, while Bia East in the Western North Region also remained above this threshold, with rates from 87.1 to 161.6. This regional variation highlights disparities in snakebite risk across Ghana’s diverse ecological zones.

In the Eastern Region, Nsawam Adoagyiri reached 200.8 in 2021—the second-highest district-level value recorded nationally that year—and recorded rates ranging from 33.2 to 104.1 in the other years. Fanteakwa North, also in the Eastern Region, reported incidences ranging from 62.3 to 174.1, with its 2025 value the highest that year across all the districts. In the Ashanti Region, Asutifi South had incidences ranging from 25.5 to 88.5, while Bosome Freho varied from 45.7 to 79.3. In the Central Region, Upper Denkyira East ranged from 27.3 to 83.4, and Twifo Hemang Lower Denkyira varied from 29.2 to 98.7. The Western Region saw Wassa Amenfi Central ranging from 41.2 to 108.1 and Wassa East from 58.0 to 100.1.

In the Eastern Region, Akyemansa reported rates ranging from 30.4 to 85.6, while in the Bono East Region, Pru West reported rates ranging from 41.5 to 84.7. In the Upper East Region, Builsa South ranged from 49.4 to 142.4, and Garu ranged from 50.1 to 139.3. This distribution of moderate-to-high incidence across various agro-ecological zones aligns with the agrarian exposure pathway discussed in Section 4. In the Greater Accra Region, Ayawaso East recorded 0.0 for all six years, while Ablekuma West recorded values from 0.0 to 2.0, and Asokore Mampong from 0.0 to 2.3 during the same period. The same pattern was observed in urbanized districts elsewhere: Kumasi in the Ashanti Region had rates ranging from 3.0 to 19.5, Sekondi Takoradi in the Western Region ranged from 3.5 to 15.6, and Suame, also in the Ashanti Region, varied from 0.0 to 6.6. Notably, Nanton in the Northern Region recorded 0.0 in five out of six years and 1.8 in the remaining year, despite being located within a predominantly high-incidence area, thus consistently remaining in the lowest national incidence category throughout the period.

#### 3.3.2 Relative Risk Mapping

Fig. 3 illustrates the posterior mean relative risk (RR) of snakebite envenoming across Ghana’s 261 districts for each year from 2020 to 2025, aiming to inform public health strategies (see Table A2 in the Appendix). This data was smoothed using the Bayesian spatio-temporal BYM2 model and mapped on a diverging scale centered around unity. The resulting surface reveals a nationally distributed risk profile, where credibly elevated risk (RR *>* 1, with the lower credible bound exceeding unity) recurs across multiple regions.

**Fig. 3.**
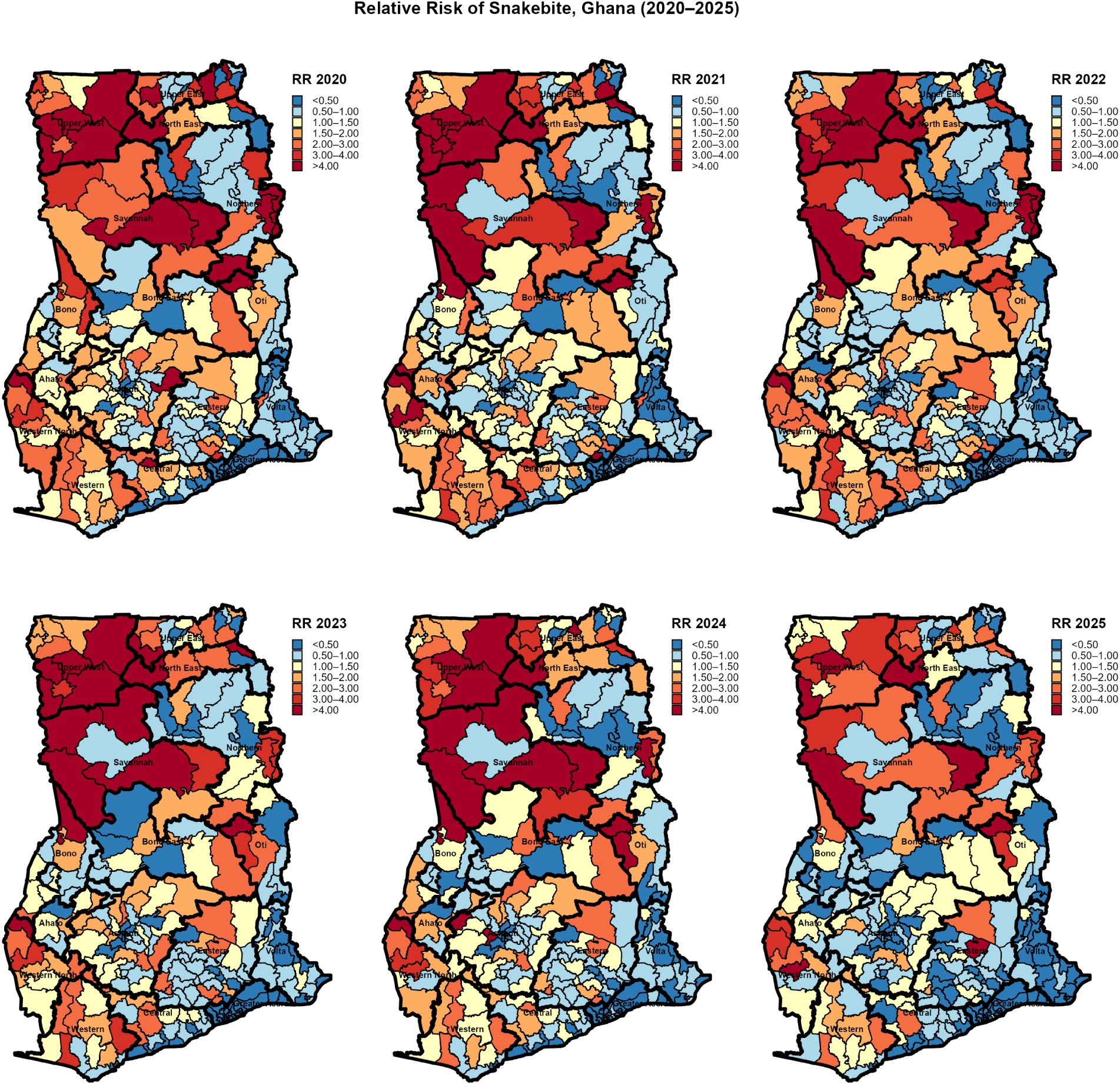
Relative risk of snakebite across the 261 districts of Ghana, by year (2020–2025), estimated from the Bayesian spatio-temporal BYM2 model. RR expresses each district’s risk relative to the national average: RR = 1 (pale) denotes average risk; values below 1 (blue) indicate lower-than-average risk; and values above 1 (red) indicate elevated risk. Estimates are smoothed by borrowing strength across neighboring districts and time periods.

Mamprugu Moagduri in the North East Region exhibited the highest RR nationally from 2020 to 2025, with values of 5.10 (95% CrI: 2.86–8.56), 8.47 (5.28–13.00), 13.31 (8.97–19.08), 11.67 (7.70–17.02), 7.30 (4.41–11.46), and 6.10 (3.59–9.79), peaking in 2022. The overall period estimate for this region was 8.69 (5.50–13.20). Daffiama Bussie Issa in the Upper West Region recorded a range from 5.70 to 9.87 during the same period, peaking in 2021, with an overall estimate of 7.48 (3.92–13.10). North East Gonja in the Savannah Region ranged from 3.42 to 7.88, peaking in 2020, with an overall estimate of 5.20 (2.45–9.84). Wa East and Wa West, both in the Upper West Region, consistently maintained an RR above 4 for five of the six years, with Wa East showing a range from 2.89 to 7.05 (overall 5.57, 3.45–8.61) and Wa West ranging from 3.91 to 6.48 (overall 5.24, 3.13–8.33). Bole in the Savannah Region increased from an RR of 1.65 (0.76–3.17, not credibly elevated) in 2020 to 7.37 (5.06–10.46) in 2024, before declining to 4.65 (2.91–7.12) in 2025. Several districts outside these regions also showed comparably high, credibly elevated RR throughout the study period, underscoring the need for focused public health efforts. Bia East in the Western North Region ranged from 3.35 to 5.41, peaking in 2020, with an overall estimate of 4.61 (2.30–8.33). Krachi Nchumuru in the Oti Region ranged from 3.57 to 5.24 (overall 4.39, 2.41–7.42), with some of the narrowest credible intervals, indicating high posterior certainty.

Banda in the Bono Region rose from an RR of 3.09 (1.08–7.05) in 2020 to 5.42 (2.37–10.77) in 2023, with an overall estimate of 4.15 (1.71–8.64), before falling to 2.42 (0.84–5.56) in 2025. Nsawam Adoagyiri in the Eastern Region achieved an RR of 6.33 (4.45–8.81) in 2021, its highest annual value, while recording a range of 1.44-3.41 in other years (overall: 3.31, 2.05–5.11). Fanteakwa North in the Eastern Region surged to an RR of 6.16 (4.35–8.94) in 2025, the highest recorded RR among all 261 districts that year, following a range from 2.34 to 2.89 in the previous five years (overall RR of 3.16, CrI: 1.61–5.86). Additionally, a broad intermediate stratum with credible intervals excluding unity in most years extended across the Ashanti, Bono, Bono East, Central, and Western Regions. Asutifi South in the Ahafo Region reported an RR ranging from 1.23 to 2.77 (overall 2.10). Upper Denkyira East and Upper Denkyira West in the Central Region recorded ranges from 1.24 to 2.69 and 1.59 to 2.55, respectively (overall 2.09 and 2.21). Wassa Amenfi West and Wassa East in the Western Region ranged from 1.07 to 2.80 and 2.25 to 3.23, respectively (overall 2.10 and 2.61). Ejura Sekyedumase in the Ashanti Region ranged from 1.31 to 2.07 (overall 1.64). This distribution reflects a widespread, credibly above-average background risk extending across multiple regions. Conversely, the lowest RR values, often below 0.10 with credible intervals entirely beneath unity, were concentrated in urban districts, regardless of region. Ablekuma Central, Ayawaso Central, and Ayawaso East in the Greater Accra Region recorded RR values between 0.02 and 0.04 every year from 2020 to 2025. Suame in the Ashanti Region saw a range from 0.08 to 0.14 during the same period, while Kumasi in the Ashanti Region ranged from 0.18 to 0.26 from 2020 to 2024 before rising to 0.64 in 2025, its highest recorded value. Nanton in the Northern Region sustained RR values between 0.05 and 0.07 throughout the study period, indicating that both low- and high-risk areas are influenced more by urbanization than by regional boundaries alone.

#### 3.3.3 Exceeding Probability Mapping

Fig. 4 illustrates the posterior exceedance probability, *P* (RR *>* 1), across Ghana’s 261 districts for each year from 2020 to 2025 (refer to Table A3 in the Appendix). This figure highlights districts where elevated snakebite risk is not only estimated but also statistically credible. Districts with *P >* 0.95 are classified as credible high-risk hotspots. Mamprugu Moagduri (North East Region) recorded a probability of *P* = 1.000 in both 2022 and 2023, maintaining a level above 0.92 in each year from 2020 to 2025, resulting in an overall probability of 0.972. North East Gonja (Savannah Region) achieved *P* = 1.000 in 2020 and remained above 0.96 from 2021 to 2024, with an overall value of 0.973. Similarly, Central Gonja (Savannah Region) reached *P* = 1.000 in 2020, concluding with an overall probability of 0.961. Wa West (Upper West Region) also attained *P* = 0.999 in three years: 2021, 2022, and 2023, resulting in an overall probability of 0.974. Wa East (Upper West Region) exceeded *P* = 0.99 in multiple years, achieving an overall probability of 0.973. Daffiama Bussie Issa (Upper West Region) reached *P* = 1.000 in 2021 and maintained an overall exceedance probability of 0.988 from 2020 to 2025. Sissala East and Bole (Savannah Region) both recorded *P* = 1.000 in 2023, with overall probabilities of 0.951 and 0.901, respectively. Bia East (Western North Region) reached *P* = 1.000 in 2023 and remained above 0.90 throughout the other years from 2020 to 2025, achieving an overall probability of 0.979. Krachi Nchumuru (Oti Region) maintained a probability between 0.951 and 0.990 from 2020 to 2024, declining to 0.873 in 2025, with an overall value of 0.961. Wassa Amenfi Central (Western Region) recorded *P* = 0.995 in 2022, resulting in an overall probability of 0.911. Ellembelle (Western Region) sustained a probability between 0.921 and 0.992 from 2020 to 2024, then declined to 0.887 in 2025, giving an overall value of 0.950.

**Fig. 4.**
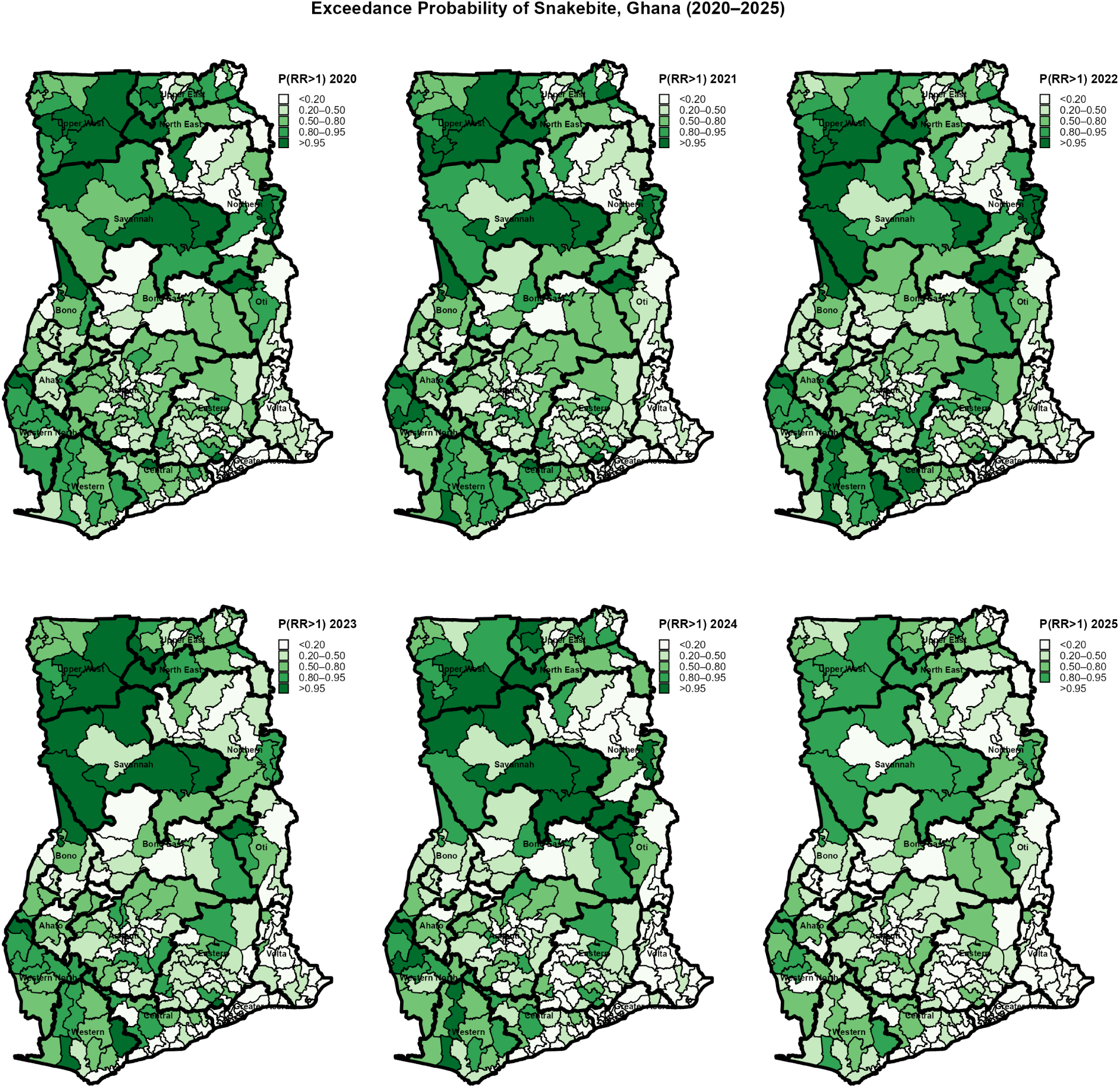
Posterior exceedance probability of snakebite relative risk, P(RR *>* 1), across the 261 districts of Ghana, by year (2020–2025), from the Bayesian spatio-temporal model. The exceedance probability quantifies the posterior certainty that a district’s RR exceeds the national average; values approaching 1 (dark green) indicate strong evidence of elevated risk, values near 0 indicate strong evidence of below-average risk, and values near 0.5 indicate uncertainty. Districts with an exceedance probability above 0.95 were classified as credible high-risk hotspots.

In the Eastern Region, Nsawam Adoagyiri reached *P* = 0.997 in 2023 and remained above 0.90 in five of the six years from 2020 to 2025, resulting in an overall probability of 0.928. Fanteakwa North sustained a probability between 0.831 and 0.940 from 2020 to 2024, declining to 0.669 in 2025, with an overall value of 0.859. Banda (Bono Region) maintained a probability between 0.913 and 0.991 from 2020 to 2024, falling to 0.807 in 2025, with an overall value of 0.937. Juaboso (Western North Region) and Wassa East (Western Region) both had overall exceedance probabilities above 0.93, at 0.936 and 0.938, respectively. A broad intermediate stratum, with exceedance probabilities generally between 0.5 and 0.9, extended across the Ashanti, Central, Bono, and Eastern Regions. Bosome Freho (Ashanti Region) had values ranging from 0.714 to 0.916 from 2020 to 2025, Assin South (Central Region) ranged from 0.791 to 0.963, Akyemansa (Eastern Region) varied from 0.493 to 0.909, and Kwahu Afram Plains South (Eastern Region) ranged from 0.717 to 0.912, indicating a moderately confident elevation in risk distributed across multiple regions. Exceedance probabilities at or near zero were not confined to a single region but were found in urbanized districts nationwide. In the Greater Accra Region, Ablekuma Central, Ablekuma West, Ayawaso Central, Ayawaso East, Krowor, Tema, and Tema West all recorded *P* = 0.000 in every year from 2020 to 2025. Kumasi (Ashanti Region) did not exceed *P* = 0.091 in any year (with an overall probability of 0.016), Suame (Ashanti Region) remained at or near 0.000. Sekondi Takoradi (Western Region) did not exceed *P* = 0.055 in any year (with an overall value of 0.020). Tempane (Upper East Region) and Nanton (Northern Region) both reported exceedance probabilities of essentially zero each year (overall 0.000 for both), illustrating that low statistical certainty of elevated risk, like high certainty, closely tracks urbanization rather than regional location.

#### 3.3.4 Hotspot Mapping

Fig. 5 illustrates the spatial clustering identified by the global Moran’s *I* statistic, highlighting key patterns to enhance understanding. Each district is categorized by local Moran’s *I* into High-High (H-H) hotspots, Low-Low (L-L) coldspots, and spatial outliers (High-Low, Low-High) for the years 2020 to 2025. The largest and most consistent H-H hotspot cluster is found in the Upper West Region. Here, Daffiama Bussie Issa, Jirapa, Nadowli Kaleo, Sissala East, Wa, Wa East, and Wa West were classified as H-H every year from 2020 to 2025 without interruption. Sissala West joined this cluster starting in 2021, having been classified as Low-High in 2020 before switching to H-H in each subsequent year. In the Savannah Region, North Gonja and Sawla Tuna Kalba were classified as H-H in all six years. Central Gonja achieved H-H status in five out of six years, with the only interruption occurring in 2022. East Gonja was classified as H-H in four of six years, with interruptions in 2022 and 2023, while Bole entered the H-H cluster intermittently in 2021, 2023, and 2024. The Upper East Region saw Builsa South and Kasena Nankana West classified as H-H every year from 2020 to 2025, while Garu and Nabdam achieved H-H status only in 2020. In the North East Region, Mamprugu Moagduri maintained H-H status in all six years, and West Mamprusi joined the cluster from 2021 onward.

**Fig. 5.**
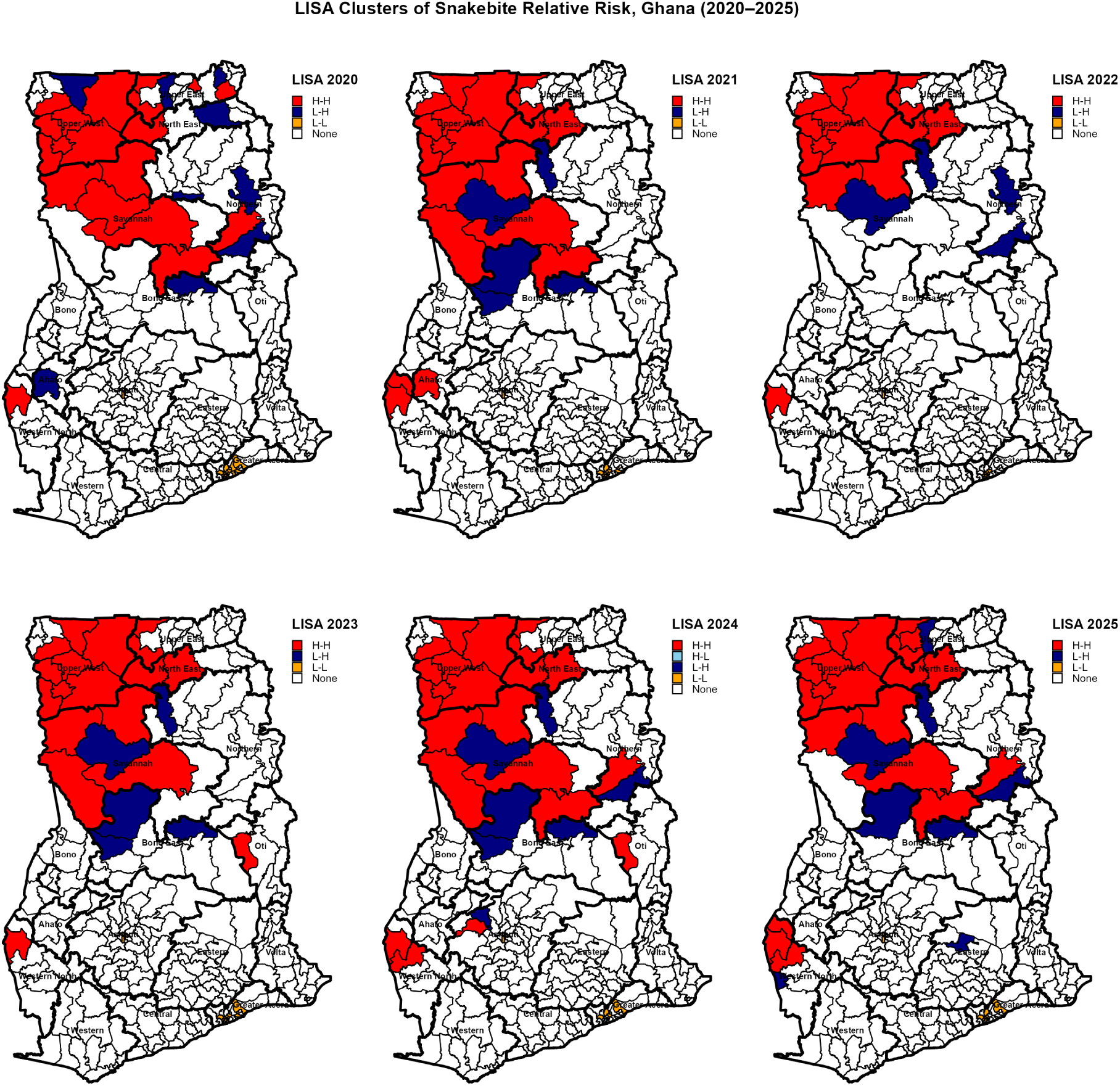
Local Indicators of Spatial Association cluster maps of snakebite relative risk by district, Ghana, 2020–2025. Districts are classified as High-High (H-H, red), High-Low (H-L, blue), Low-High (L-H, navy), or Low-Low (L-L, orange) clusters based on statistically significant local spatial autocorrelation (*p ≤* 0.05); districts with non-significant associations are shown in white (None). H-H clusters indicate districts with high relative risk surrounded by neighboring districts of similarly high risk, while L-L clusters indicate low-risk districts surrounded by low-risk neighbors.

Significant H-H clustering was not limited to these four regions, underscoring the prevalence of high-risk zones. In the Western North Region, Bia West was classified as H-H every year from 2020 to 2025, demonstrating the same temporal consistency as the strongest districts in Upper West. Bia East attained H-H status in 2021 and 2025, while Juaboso achieved it in 2024 and 2025. In the Oti Region, Krachi West entered the H-H cluster in 2023 and 2024. In the Northern Region, Nanumba North registered H-H clustering in 2020, 2024, and 2025. These findings indicate significant spatial clustering of elevated risk in at least six regions: Upper West, Savannah, North East, Upper East, Western North, and Oti. Low-High spatial outliers, representing low-risk districts adjacent to high-risk neighbors, were found across several regions. For example, West Gonja in the Savannah Region registered Low-High status in five out of six years from 2021 to 2025, despite bordering persistently high-risk Gonja districts. Kumbungu in the Northern Region exhibited a similar pattern, registering Low-High in five out of six years, also from 2021 to 2025. In the Bono East Region, two districts contributed to this pattern: Kintampo North registered Low-High in four years (2021, 2023, 2024, and 2025), while Pru registered Low-High in five out of six years (2020, 2021, 2023, 2024, and 2025). The Low-Low coldspot cluster, indicating low-risk districts surrounded by similarly low-risk neighbors, was concentrated in the Greater Accra Region. Ablekuma North, Ayawaso Central, Ayawaso West, Ga Central, and Okai Koi North each registered Low-Low classification in four or more of the six years from 2020 to 2025. Accra itself was classified as Low-Low in five years, with only a single High-Low outlier year in 2024. Kumasi in the Ashanti Region registered Low-Low status every year from 2020 to 2025, providing reassurance about the stability of low-risk areas outside the capital, which is vital for regional planning and public confidence.

#### 3.3.5 Geographic Accessibility to Snakebite Treatment Facilities

Fig. 6 illustrates the straight-line (Euclidean) distance from each district centroid to the nearest snakebite treatment health facility across Ghana’s 261 districts. Highlighting these geographic disparities can guide policymakers to focus on regions with the greatest access challenges, especially where high-risk districts are far from facilities. The distance to the nearest health facility varies significantly, ranging from 0.1 km in Kumasi (Ashanti Region) to 86.7 km in North Gonja (Savannah Region). The national median distance is 12.8 km, while the mean distance is 17.9 km. Recognizing these disparities can motivate policymakers to prioritize regions with the greatest access challenges, especially where high-risk districts are far from facilities. Access to snakebite treatment in formal health facilities varies widely across regions. The Savannah Region records the highest regional mean distance at 54.7 km, which includes North Gonja (86.7 km), West Gonja (82.7 km), Central Gonja (56.4 km), Sawla Tuna Kalba (51.5 km), Bole (50.8 km), North East Gonja (30.4 km), and East Gonja (24.3 km). The North East Region follows closely, with a regional mean distance of 46.0 km, comprising Yunyoo Nasuan (60.3 km), Bunkpurugu Nakpanduri (58.2 km), Chereponi (54.0 km), Mamprugu Moagduri (52.5 km), East Mamprusi (43.4 km), and West Mamprusi (7.4 km).

**Fig. 6.**
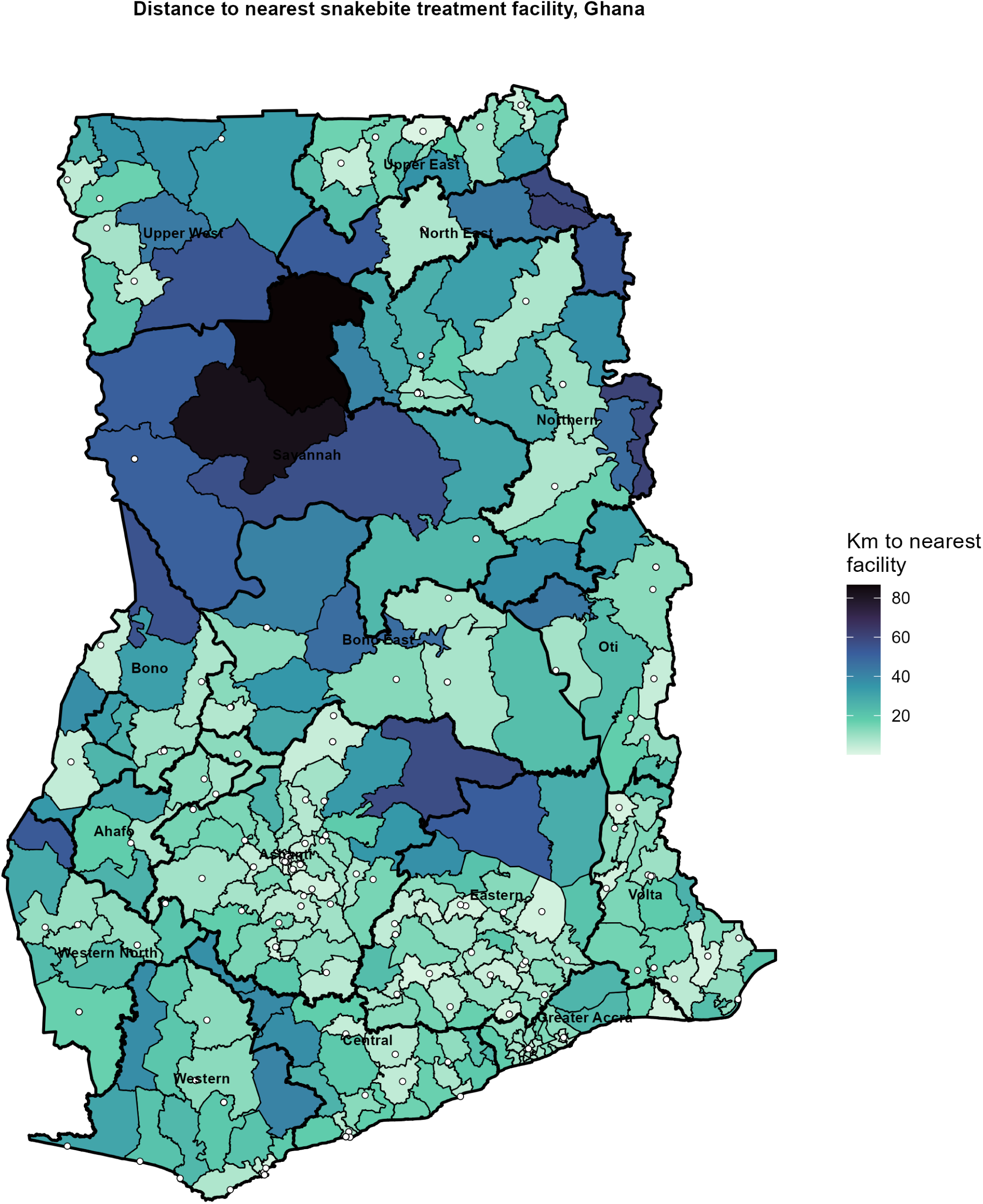
Geographic accessibility to snakebite treatment facilities, Ghana. District shading represents the straight-line (Euclidean) distance in kilometers from each district centroid to the nearest geocoded health facility; white circles denote the locations of individual treatment facilities. Darker shading indicates greater distance and correspondingly poorer facility access. Distances were computed in a projected coordinate system (UTM Zone 30N) to ensure metric accuracy.

The Northern Region has a regional mean of 26.1 km, and the Upper West Region has 25.8 km, both significantly above the national mean of 17.9 km. This highlights the need for health officials to address geographic disparities, especially in high-risk districts far from treatment facilities. In contrast, the Greater Accra Region has the lowest regional mean distance at 9.4 km, followed by the Ashanti Region at 11.5 km and the Eastern Region at 12.6 km. Some of the highest-risk districts identified in Figs. 3, 4, and 5 are also among those farthest from treatment facilities. For instance, Wa East in the Upper West Region records 54.3 km; Mamprugu Moagduri in the North East Region records 52.5 km; Bole in the Savannah Region records 50.8 km; and Sissala East in the Upper West Region records 33.3 km. Daffiama Bussie Issa, also in the Upper West Region, records 43.3 km. North East Gonja in the Savannah Region records 30.4 km.

Access to treatment varies considerably across hotspot districts identified by the incidence, relative risk, exceedance probability, and LISA analyses. For example, Banda in the Bono Region records 55.4 km; Bia East in the Western North Region records 53.2 km; Krachi Nchumuru in the Oti Region records 43.9 km; and Wassa East in the Western Region records 40.6 km. This uneven access underscores the need for targeted interventions to improve geographic coverage. In contrast, other hotspot districts are relatively well-served. For instance, Nsawam Adoagyiri in the Eastern Region is only 2.1 km away, despite being one of the districts with the highest probability of exceedance nationally. Fanteakwa North, also in the Eastern Region and identified as the district with the highest relative risk in 2025, records 12.6 km. Other well-served districts include Krachi West in the Oti Region (8.7 km), Nanumba North in the Northern Region (7.3 km), Juaboso in the Western North Region (10.7 km), Wassa Amenfi Central (20.0 km), and Ellembelle (21.0 km), both in the Western Region. This discrepancy indicates that geographic access to snakebite treatment is not uniformly poor across all districts identified as statistically credible hotspots.

## 4 Discussion

This study provides the first nationwide Bayesian spatio-temporal assessment of snakebite envenoming risk across all 261 districts of Ghana using six years of routine surveillance data from Ghana’s DHIMS-2. By integrating spatial and temporal random effects with environmental covariates, hotspot detection, posterior exceedance probabilities, and geographic accessibility analyses within a unified modeling framework, the study moves beyond conventional descriptive surveillance to provide a probabilistic understanding of where snakebite risk is concentrated, how it evolves, and where health-system vulnerabilities are likely to be greatest. This comprehensive approach offers valuable insights to inform effective public health strategies and resource allocation.

Several important findings emerge from this analysis. First, snakebite risk was highly heterogeneous across Ghana, exhibiting marked spatial clustering rather than a uniform national distribution. Persistent areas of elevated risk were concentrated predominantly within the northern savanna ecological zone, particularly across districts in the Upper West (Daffiama Bussie Issa, Wa East, Wa West, and Sissala East), Savannah (Bole and the Gonja districts), and North East Regions (Mamprugu Moagduri). However, important new hotspots were also identified within the Western North (Bia East), Bono (Banda), Oti (Krachi Nchumuru), Eastern (Nsawam Adoagyiri and Fanteakwa North), and Western (Wassa East) Regions. Second, relative humidity, together with both maximum and minimum temperatures, was positively associated with snakebite risk after accounting for spatial and temporal dependence. In contrast, rainfall and NDVI were not credibly associated with district-level incidence within the fitted model. Third, multiple complementary mapping approaches, including model-based relative risks, posterior exceedance probabilities, and LISA, identified remarkably consistent hotspot patterns, providing strong evidence that the observed geographic heterogeneity reflects genuine epidemiological variation rather than random fluctuations in surveillance data. Finally, the integration of healthcare accessibility revealed an important spatial mismatch: several districts, including Wa East, Mamprugu Moagduri, Daffiama Bussie Issa, Sissala East, North East Gonja, Banda, Bia East, Krachi Nchumuru, and Wassa East, with the greatest estimated snakebite burden, also had comparatively poor geographic access to treatment facilities. Collectively, these findings provide an evidence base for geographically targeted surveillance, strategic antivenom deployment, and prioritization of health-system investments.

Taken together, this study establishes a strong epidemiological foundation for understanding the contemporary geography of snakebite in Ghana. The following sections explore the environmental and ecological mechanisms that may explain the observed spatial patterns and discuss their implications for snakebite prevention and control.

### Environmental determinants of snakebite risk

The identification of relative humidity and both maximum and minimum temperatures as significant environmental correlates of snakebite risk highlights the important influence of climatic conditions [28, 29] on the spatial ecology of snakebite in Ghana. These associations are biologically plausible because snakes are ectothermic animals [30] whose physiology, activity patterns, prey availability, and reproductive behavior are strongly influenced by ambient environmental conditions. Variations in temperature and humidity affect thermoregulation, movement, shelter selection, and foraging activity, which in turn modify the frequency of encounters between humans and snakes [29, 31–33]. Importantly, the associations observed in this study should be interpreted as population-level relationships at the district scale rather than direct causal effects at the individual level. Nevertheless, they provide valuable evidence that climatic variability helps shape the geographic distribution of snakebite risk across Ghana.

The positive association between temperature and snakebite risk is consistent with ecological theory and previous epidemiological investigations [34–36]. Warmer conditions generally increase snake activity by facilitating thermoregulation and extending periods when hunting and movement are optimal. At the same time, higher temperatures often coincide with increased agricultural activity, livestock grazing, and other outdoor occupations that increase human exposure to snake habitats. Consequently, elevated snakebite incidence may reflect the combined effects of increased snake activity and greater opportunities for human-snake contact rather than temperature acting independently. Similar relationships have been reported across several snakebite-endemic settings, in which temperature consistently emerges as one of the strongest climatic predictors of snakebite occurrence [35–39]. The present findings therefore reinforce growing evidence that temperature is an important environmental determinant of snakebite risk across diverse ecological settings.

Relative humidity demonstrated the strongest environmental association in the fitted model. Humid conditions may influence the occurrence of snakebite through several complementary mechanisms. Increased humidity can improve habitat suitability for many medically important snake species by reducing desiccation risk and maintaining favorable microclimatic conditions within vegetation, burrows, and agricultural landscapes. Humidity may also indirectly influence prey abundance, thereby increasing snake foraging activity in areas where humans work or reside. From a behavioral perspective, periods of high humidity often coincide with farming activities requiring prolonged outdoor exposure, particularly in rain-fed agricultural systems that dominate many high-risk districts identified in this study. The observed association therefore likely reflects the combined influence of ecological suitability and human occupational exposure rather than a single environmental pathway. Our findings are consistent with the broader international literature demonstrating that humidity is an important predictor of snakebite risk [19, 30, 35, 40]. The concordance among studies conducted in markedly different ecological regions suggests that humidity may be a robust, large-scale determinant of snakebite risk, despite differences in snake fauna, climatic regimes, and human populations.

While rainfall was not statistically associated with snakebite risk in this analysis, this does not mean precipitation plays no ecological role in snakebite occurrences. Rainfall affects snake ecology through various pathways over different time scales [20, 30]. Rainfall can promote vegetation growth and increase the abundance and activity of prey species, thereby enhancing habitat suitability and foraging opportunities for snakes [41, 42]. Evidence from Ghana shows that flooding can increase snakebite cases as snakes move into human areas after heavy rain [43]. Additionally, the lack of a significant association between NDVI and snakebite risk should be interpreted carefully. NDVI measures vegetation greenness [44] rather than habitat quality or prey availability, and district-level averages can mask local land-cover variations that influence snake populations. This does not mean vegetation is not important; it simply indicates that NDVI alone does not capture the full ecological picture.

### Spatial heterogeneity and ecological interpretation of hotspot persistence and emergence

A defining feature of this study is the pronounced spatial heterogeneity of snakebite risk across Ghana. The present findings therefore reinforce the importance of analyzing snakebite within an explicitly spatial framework rather than relying solely on aggregated regional or national incidence estimates [45], which can conceal considerable within-region heterogeneity. Rather than being randomly distributed, snakebite exhibited persistent geographic clustering, with elevated risk concentrated within distinct ecological landscapes that remained relatively stable throughout the six-year study period. In particular, the northern savanna ecological zone consistently represented the principal focus of snakebite risk. Districts within the Upper West (Daffiama Bussie Issa, Wa East, Wa West, and Sissala East), Savannah (Bole and the Gonja districts), and North East (Mamprugu Moagduri) Regions repeatedly demonstrated elevated relative risks. They formed contiguous areas of increased disease burden across successive years. This observation is consistent with previous studies from northern Ghana that documented disproportionately high snakebite incidence in predominantly agricultural communities [6, 41]. Our findings extend these regional observations by demonstrating that the northern corridor forms part of a broader national pattern that remains evident after accounting for differences in population size, spatial dependence, temporal variation, and environmental covariates.

Although the northern savanna has remained the predominant hotspot for snakebites throughout the study period, another significant finding was the identification of persistently high-risk districts outside this traditional endemic zone. Districts in the Western North (Bia East), Bono (Banda), Oti (Krachi Nchumuru), Eastern (Nsawam Adoagyiri and Fanteakwa North), and Western (Wassa East) Regions consistently exhibited elevated relative risks and credible hotspot classifications, despite being situated in markedly different ecological settings. These findings suggest that a simple north-south dichotomy cannot accurately characterize Ghana’s snakebite burden [45]. Instead, various epidemiological landscapes appear to contribute to the national burden, each likely driven by distinct combinations of ecological, occupational, and socioeconomic factors [7, 41, 46].

The emergence of high-risk districts in forest and forest-transition zones carries significant ecological implications. Unlike the northern savanna, these areas are characterized by dense vegetation, cocoa cultivation, mixed farming systems, forest reserves, and expanding human settlements. Such environments support diverse snake communities, including West African Carpet Viper (*Echis ocellatus*), West African Green Tree Viper (*Atheris chloroechis*), West African Gaboon Viper (*Bitis rhinoceros*), Variable Burrowing Asp (*Atractaspis irregularis*), and the West African Gaboon Viper (*Bitis rhinoceros*) [7], and expose populations to occupational hazards not commonly encountered in open savanna ecosystems. Consequently, the elevated risk of snakebites in these districts likely reflects alternative ecological pathways rather than merely an extension of the northern transmission pattern. The identification of persistent hotspots in both savanna and forest-transition environments underscores the need for snakebite prevention strategies to be adapted to local ecological contexts rather than assuming a uniform national risk profile.

Notably, Fanteakwa North emerged as the district with the highest estimated relative risk nationally in 2025. This observation demonstrates that snakebite risk is dynamic and can change significantly over relatively short time periods. Although the present study was not designed to determine the precise drivers of this increase, several mechanisms may plausibly contribute, including environmental changes, agricultural expansion, land-use changes, fluctuations in snake populations, shifts in healthcare-seeking behavior, or improvements in disease reporting. Regardless of the underlying cause, this finding illustrates that relying solely on historically recognized hotspot districts may overlook newly emerging areas of increased risk. Therefore, dynamic surveillance systems that can periodically update spatial risk estimates are essential for maintaining effective snakebite prevention and control programs in Ghana.

The identification of emerging hotspots also has broader implications for climate adaptation and environmental change. Throughout tropical regions, ongoing changes in rainfall patterns, temperature, vegetation structure, and land use are increasingly recognized as factors that affect wildlife distributions and human-wildlife interactions. As a result, snakebite risk is unlikely to remain geographically static over the coming decades. Instead, ecological transitions may alter both the suitability of snake habitats and patterns of human exposure, leading to the emergence of new high-risk districts while potentially reducing risk in others. The temporal variation observed in this study provides empirical support for this dynamic perspective, highlighting the importance of integrating routine surveillance with environmental monitoring to identify evolving risk patterns before they become entrenched.

Additionally, the consistently low-risk districts found in highly urbanized areas, such as Greater Accra (Ablekuma North, Ayawaso Central, Ayawaso West, Ga Central, and Okai Koi North) and Kumasi, provide valuable insights into snakebite risks. Urban environments typically offer fewer suitable habitats for many medically important venomous snakes due to factors such as habitat fragmentation, reduced natural vegetation, a lower reliance on agricultural occupations, and greater distances between human settlements and wildlife habitats. Although snakebites can occur in urban settings [47], the consistently low relative risks in these districts highlight how ecological factors and occupational exposure significantly influence disease distribution. This contrast suggests that geographic variations in snakebite risk are less about administrative boundaries and more about the interaction between environmental conditions and human activities.

Overall, these findings indicate that the spatial distribution of snakebites in Ghana should be understood as the result of multiple interacting ecological systems, rather than a single national process. Persistent hotspots reflect long-standing environmental and occupational conditions that lead to increased human-snake interactions. In contrast, emerging hotspots show that these systems are dynamic and can respond to changes in ecological and societal conditions. Recognizing both persistence and emergence is essential for designing effective surveillance systems that can identify not only areas with a high current burden of snakebites but also regions where future public health challenges may arise. This ecological perspective serves as an important foundation for interpreting the probabilistic disease maps produced by the Bayesian framework, which is discussed in the following section.

### Bayesian disease mapping, relative risk estimation, and spatial uncertainty

This study makes a significant contribution by not only identifying districts at high risk of snakebite but also quantifying the uncertainty in these estimates. Disease surveillance data collected from routine health information systems are inherently heterogeneous, with considerable variation in population size, reporting completeness, and case counts across districts [48]. While the annual reported incidence of snakebites varied widely across districts, the model pinpointed areas such as Upper West (Daffiama Bussie Issa, Wa East, Wa West, and Sissala East), Savannah (Bole and the Gonja districts), and North East (Mamprugu Moagduri), Western North (Bia East), Bono (Banda), Oti (Krachi Nchumuru), Eastern (Nsawam Adoagyiri and Fanteakwa North), and Western (Wassa East) Regions of elevated relative risk that remained stable over time. This distinction is crucial from an epidemiological perspective, as public health decisions should ideally be informed by sustained patterns of disease risk rather than by isolated fluctuations due to random variation or reporting inconsistencies. By differentiating persistent spatial patterns from random noise, Bayesian disease mapping offers a solid basis for identifying districts where intensified surveillance and interventions are most likely to yield meaningful public health benefits.

Another strength of this analysis is the use of posterior exceedance probabilities [49, 50] to complement relative risk estimation. This probabilistic approach provides much more insight than point estimates alone, as it highlights districts where the evidence strongly supports elevated risk and contrasts them with areas where high relative risks are uncertain. From a public health standpoint, exceedance probabilities provide a transparent framework for prioritizing interventions under resource constraints [50]. Health authorities often cannot implement comprehensive snakebite prevention programs across all districts at once. As a result, decisions regarding the distribution of antivenom, community education efforts, surveillance enhancements, and emergency preparedness must be explicitly prioritized. Same districts in the northern savanna ecological zone, as well as Bia East, Banda, Krachi Nchumuru, Nsawam Adoagyiri, Fanteakwa North, and Wassa East, exhibiting elevated high relative risk with posterior certainty, present the strongest case for intervention since the likelihood of misclassifying disease burden is comparatively low. In contrast, districts with moderately elevated relative risks but substantial uncertainty, such as Bosome Freho (Ashanti Region), Akyemansa and Kwahu Afram Plains South (Eastern Region), Sefwi Wiawso (Western North Region), and Talensi (Upper East Region), may require enhanced surveillance before significant resource investments are made. This distinction enables evidence-based prioritization that considers both epidemiological need and statistical confidence.

Unlike relative risk estimation, which assesses disease burden within individual districts, LISA examines whether neighboring districts show similar patterns of elevated or reduced risk. The identification of persistent high-high clusters, including Daffiama Bussie Issa, Jirapa, Nadowli Kaleo, Sissala East, Wa, Wa East, and Wa West in the Upper West Region; North Gonja and Sawla Tuna Kalba in the Savannah Region; Mamprugu Moagduri in the North East Region; Builsa South and Kasena Nankana West in the Upper East Region; and Bia West in the Western North Region across multiple years indicates that snakebite risk in Ghana is spatially organized rather than randomly distributed. This finding has important epidemiological implications, suggesting that neighboring districts often share common environmental conditions, agricultural practices, occupational exposures, and ecological characteristics that collectively contribute to higher snakebite incidence.

The ongoing local spatial clustering across successive years further indicates that the underlying factors influencing snakebite risk remain relatively stable, despite temporal fluctuations in individual district estimates. This stability presents an opportunity for coordinated public health interventions that extend beyond administrative boundaries. Instead of implementing district-specific programs in isolation, neighboring hotspot districts could be addressed through regional surveillance networks, coordinated antivenom supply chains, unified community education campaigns, and collaborative emergency referral systems. Since the ecological factors driving snakebites often cross administrative borders, interventions designed around cohesive epidemiological landscapes may prove more effective than those executed independently within each district.

### Geographic accessibility, health-system implications, and policy relevance

Integrating epidemiological risk mapping with healthcare accessibility analysis is a key strength of this study. It highlights districts where the risk of snakebite overlaps with vulnerabilities in the healthcare system, providing solid evidence to prioritize public health actions. The study revealed a significant geographic mismatch between the burden of snakebites and access to treatment facilities. Many districts, such as Wa East, Mamprugu Moagduri, Daffiama Bussie Issa, Sissala East, North East Gonja, Banda, Bia East, Krachi Nchumuru, and Wassa East, experience high rates of snakebites but have limited physical access to care. Meanwhile, geographic accessibility plays a crucial role in snakebite outcomes because delays in reaching health facilities can hinder the timely administration of effective antivenom and supportive care, thus increasing the risks of mortality, permanent disability, and other severe complications [51–53]. Our findings align with evidence from sub-Saharan Africa [54, 55], indicating that poor access to snakebite treatment is a substantial barrier to equitable healthcare. Delays in seeking medical attention have been associated with higher mortality rates, longer hospital stays, and increased reliance on traditional healers [4, 56].

The WHO recognizes equitable access to antivenom as a key objective for reducing the burden of snakebites [53]. Our results confirm that similar challenges exist in Ghana, with significant variations across districts. Importantly, not all high-burden districts have inadequate access; for example, Nsawam Adoagyiri and Fanteakwa North in the Eastern Region exhibit a high risk of snakebites while being close to treatment facilities. This distinction has important implications for resource allocation, suggesting that different strategies may be necessary based on local conditions. Areas with reasonable access may require improvements in clinical capacity and antivenom availability. Meanwhile, districts with high snakebite risk and poor accessibility will need broader health system interventions, including decentralized antivenom distribution, enhanced emergency treatment capabilities, improved referral systems, and investment in local health facilities [4, 53, 57]. This approach is likely to improve both the efficiency and equity of resource allocation. Aligning resources such as antivenom distribution and emergency transport with epidemiological risk can reduce preventable mortality and disability, closely aligning with the WHO roadmap for addressing snakebite envenoming.

### Strengths and limitations

This study has several significant strengths that enhance the robustness and relevance of its findings for public health. It represents the first nationwide Bayesian spatio-temporal assessment of snakebite envenoming across all 261 districts of Ghana, utilizing six years of routine surveillance data. By integrating various spatial and temporal factors within a single analytical framework, the study provides a comprehensive understanding of snakebite epidemiology in Ghana. The Bayesian approach employed in this research generates smoothed district-level risk estimates accompanied by measures of uncertainty. This offers a solid foundation for identifying priority intervention areas. Furthermore, the use of complementary analytical methods reinforces confidence in the identified hotspot districts, indicating they are persistent phenomena rather than mere statistical artifacts. This analysis also underscores the regions where high epidemiological risk coincides with barriers to timely treatment, enhancing the practical utility of the findings for targeted prevention efforts and resource allocation. Finally, the study demonstrates the value of routinely collected health information systems in the surveillance of neglected tropical diseases. It shows how administrative datasets can be transformed into actionable evidence using modern Bayesian methods, providing a scalable framework that can be updated with new data and applied to other environmentally influenced diseases in Ghana and similar contexts.

While this study has strengths, its limitations should be acknowledged to contextualize the findings. First, the data from the DHIMS-2 only includes cases from formal healthcare facilities, leading to underreported snakebites, especially in remote areas. Second, treatment records are based on the district of care rather than the actual snakebite location, which can result in spatial misclassification. Future systems should include georeferenced locations for better insights. Third, environmental variables were analyzed as district averages, potentially obscuring local variations. A focus on finer spatial data could improve understanding of snakebite risk factors. Lastly, the lack of detailed clinical data limits insights into snake species and treatment outcomes. Integrating surveillance with clinical and ecological data could provide a clearer picture of the interactions between snakebites and health outcomes.

### Future directions

The highlighted limitations indicate key priorities for future research. Initiating prospective community-based surveillance would help quantify the extent of underreporting and improve our understanding of the true national burden of snakebites. By incorporating higher-resolution environmental data, distributed lag and non-linear climatic models, land-use change indicators, and species distribution modeling, we could gain deeper insights into the ecological factors driving spatial variations in risk. Longitudinal studies assessing the effects of climate variability, agricultural expansion, and environmental change would enhance our understanding of the drivers behind the emergence of new hotspot districts.

From a health systems perspective, future research should focus on several key areas: travel time to treatment, referral pathways, antivenom availability, healthcare worker preparedness, treatment delays, and patient outcomes. This will help identify the most effective strategies for reducing mortality and disability.

The analytical framework developed in this study has broader applicability to snakebite-endemic countries seeking to enhance evidence-informed public health planning. The integration of Bayesian disease mapping, environmental modeling, hotspot detection, uncertainty quantification, and healthcare accessibility analysis offers a transferable methodology for pinpointing priority areas, monitoring changes in disease risk patterns, and evaluating geographic equity in access to care. As nations increasingly improve routine surveillance systems for neglected tropical diseases, incorporating advanced spatial epidemiological methods into national surveillance programs will become vital for translating data into effective public health actions.

## Conclusion

This study demonstrates that significant spatial heterogeneity, dynamic temporal patterns, and substantial inequities in geographic access to treatment characterize snakebite envenoming in Ghana. By combining modern Bayesian spatio-temporal modeling with environmental and health system analyses, this research provides a solid evidence base for identifying districts where prevention, surveillance, and clinical resources can achieve the greatest public health benefit. The findings advocate for a shift from reactive, uniformly distributed interventions toward geographically targeted, data-driven strategies that account for both epidemiological risk and health system vulnerability. This approach aligns with the WHO strategy for halving snakebite envenoming by 20230. It offers a crucial foundation for reducing preventable mortality and disability while enhancing the resilience of Ghana’s health system.

## Funding

This research did not receive any specific grant from funding agencies.

### Ethics Statement

As part of the Integrated Disease Surveillance and Response, snakebite cases are routinely reported in DHIMS-2. Consequently, the data used in this study were secondary data. We confirm that no personally identifiable or confidential information was accessed or used at any stage of the analysis.

### Data Availability Statement

The data that support the findings of this study are available from the corresponding author, upon reasonable request

## Appendix A Additional spatial results

### District-level annual snakebite incidence per 100,000 population, relative risk estimates, and cluster classification in Ghana, 2020–2025

**Table A1:**
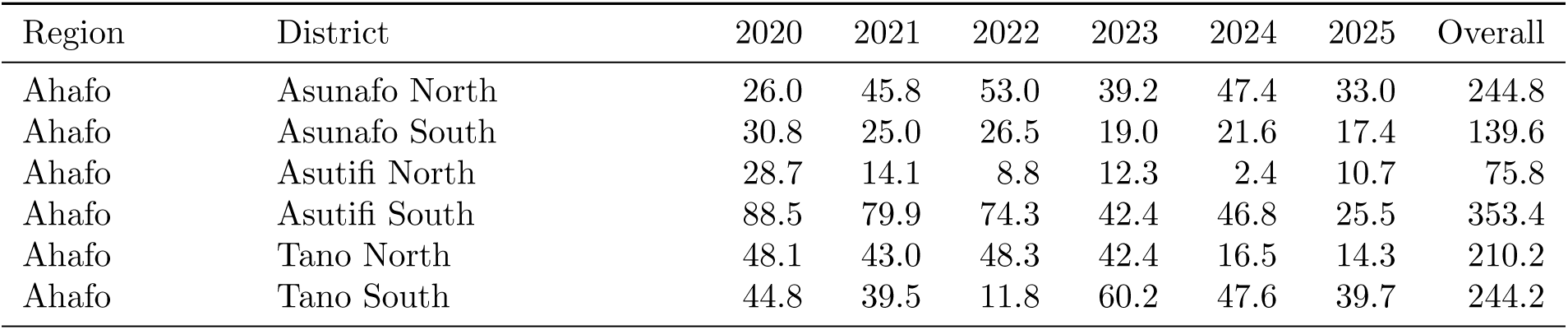
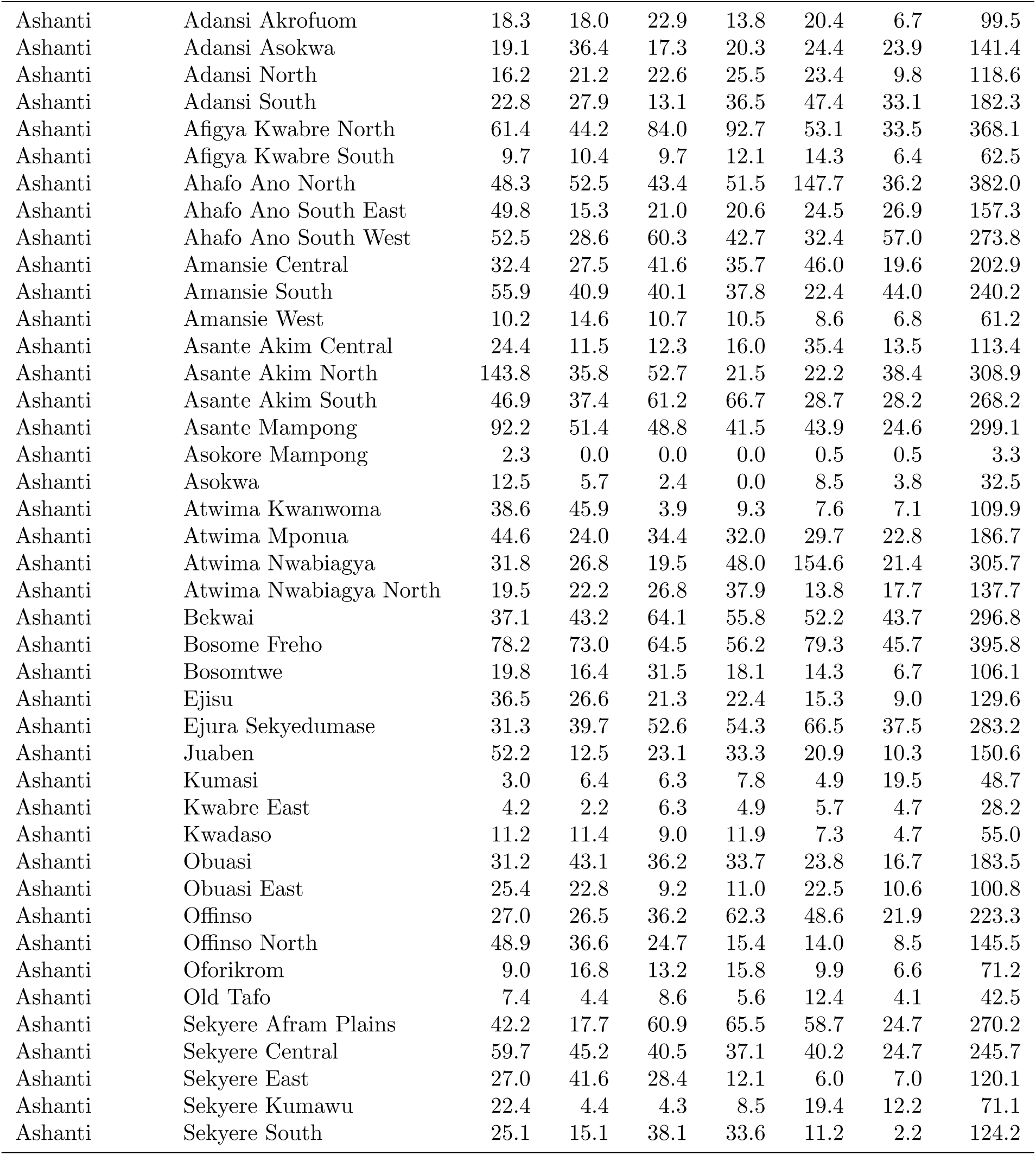
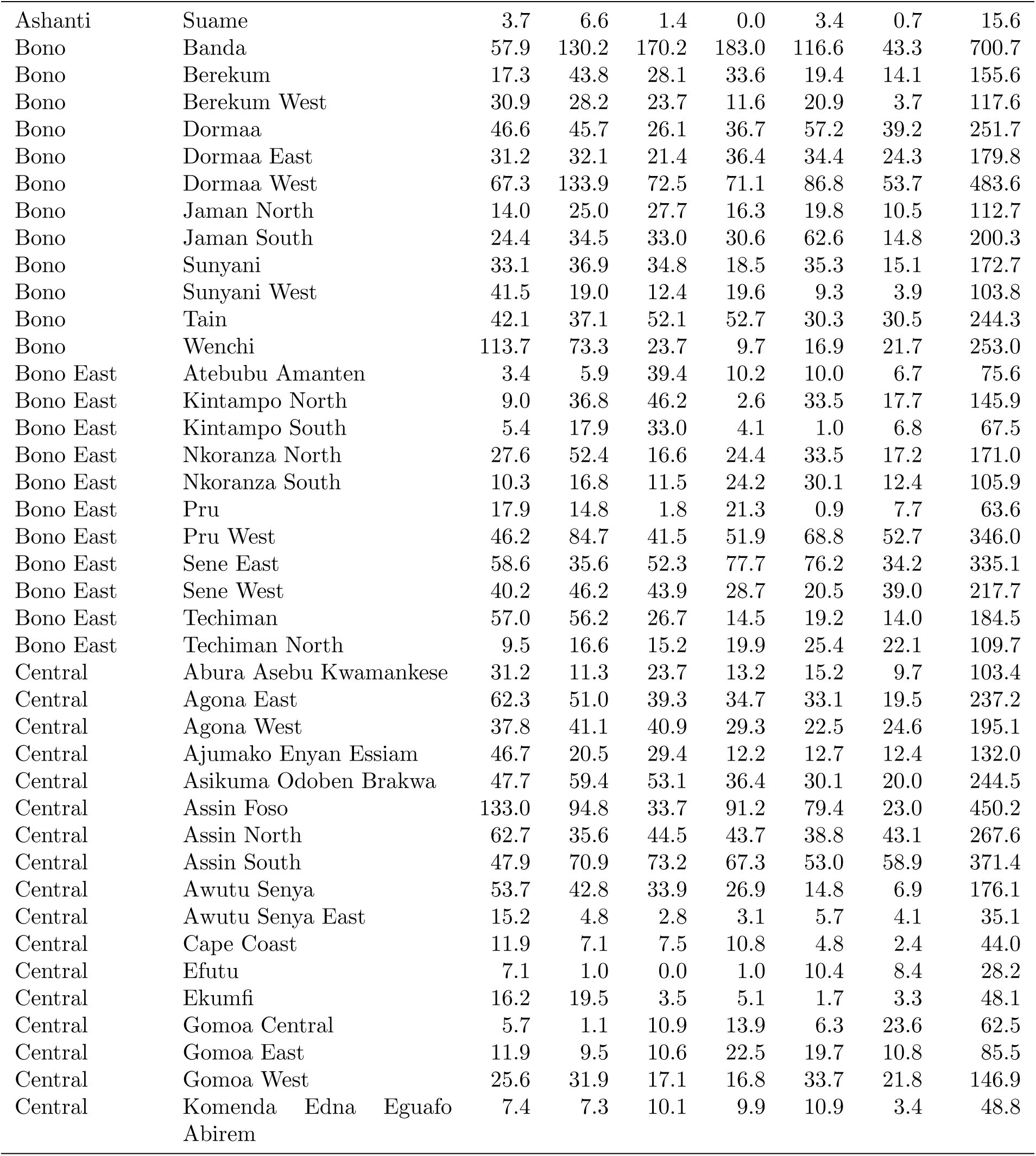
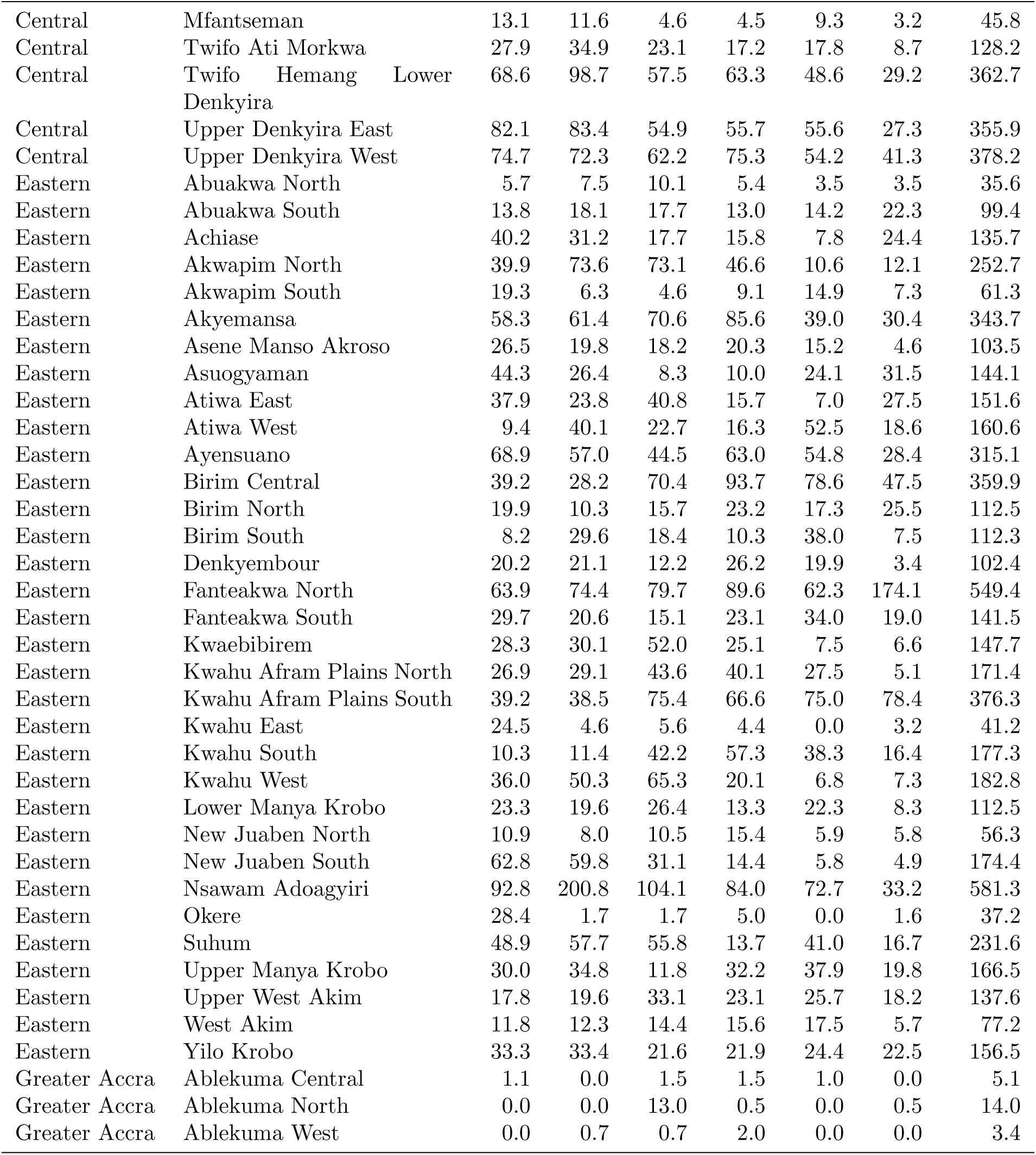
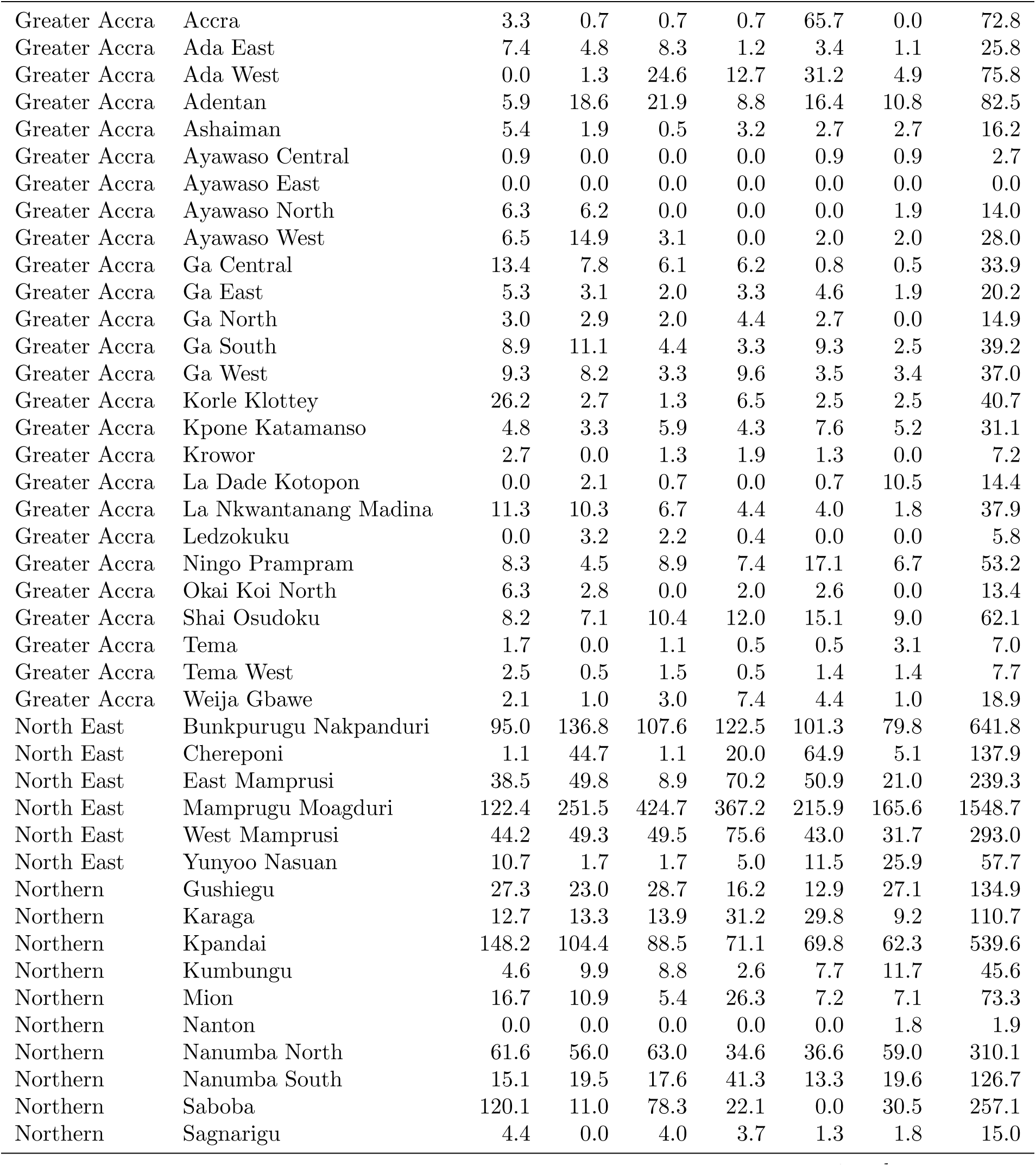
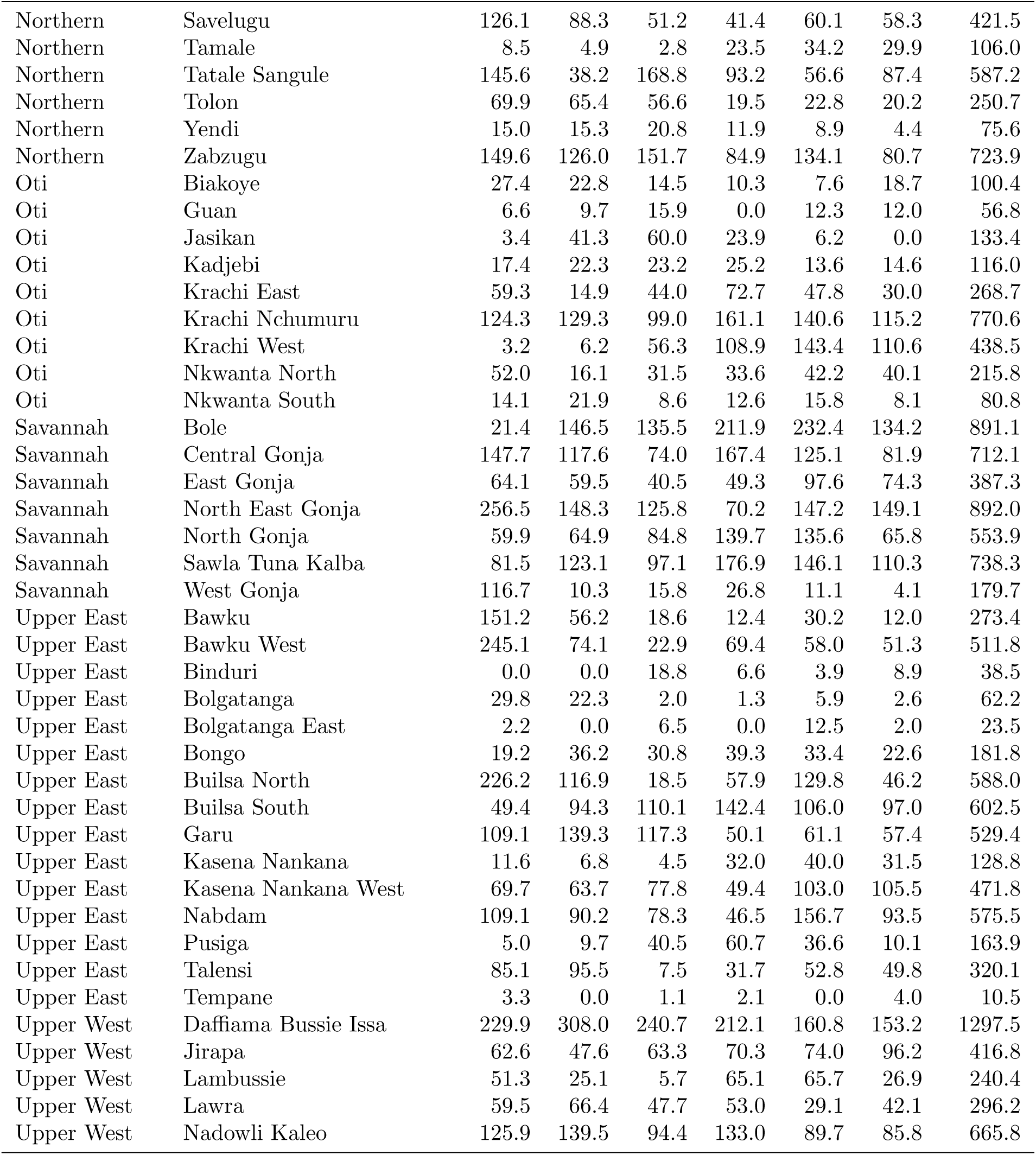
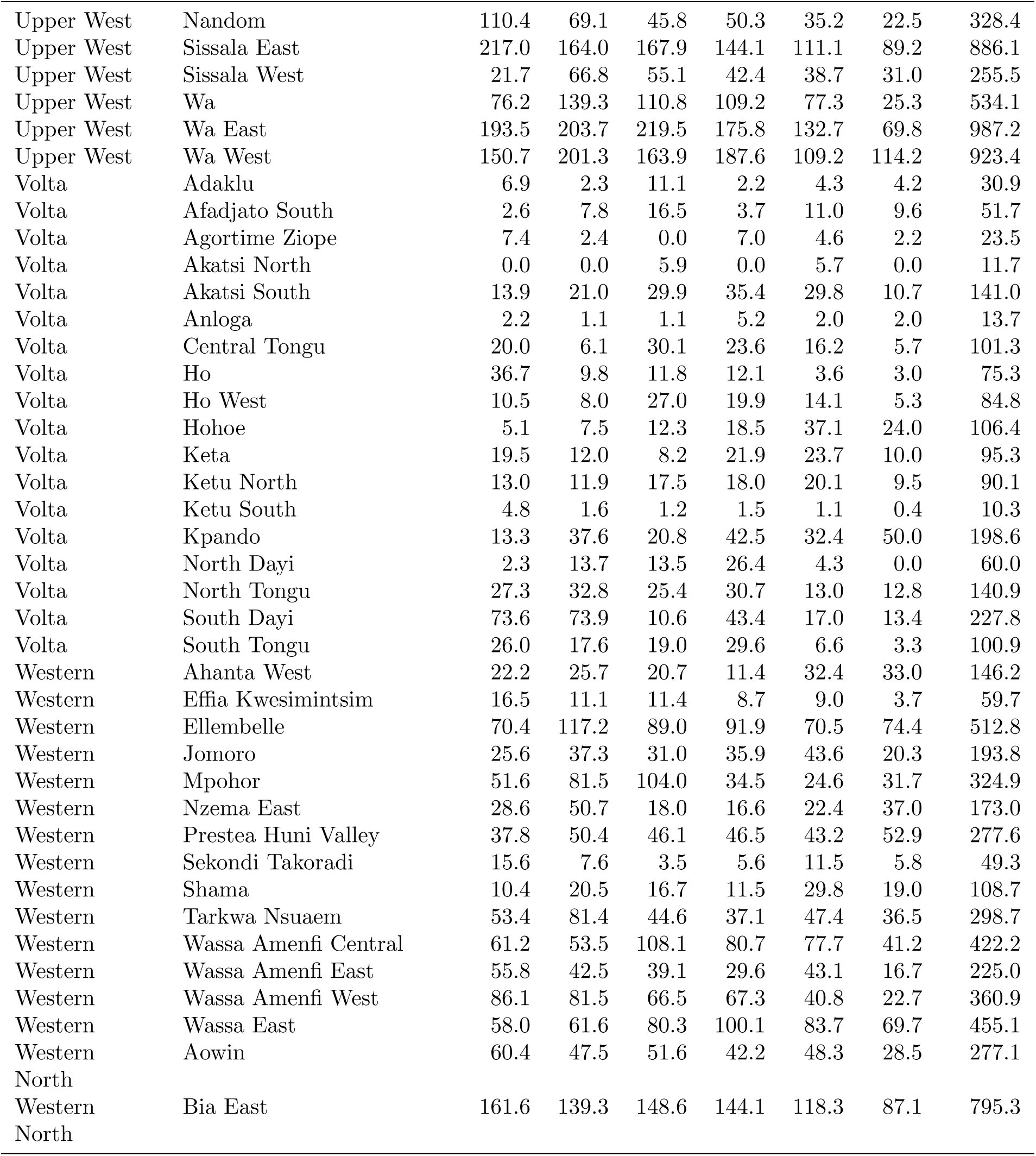
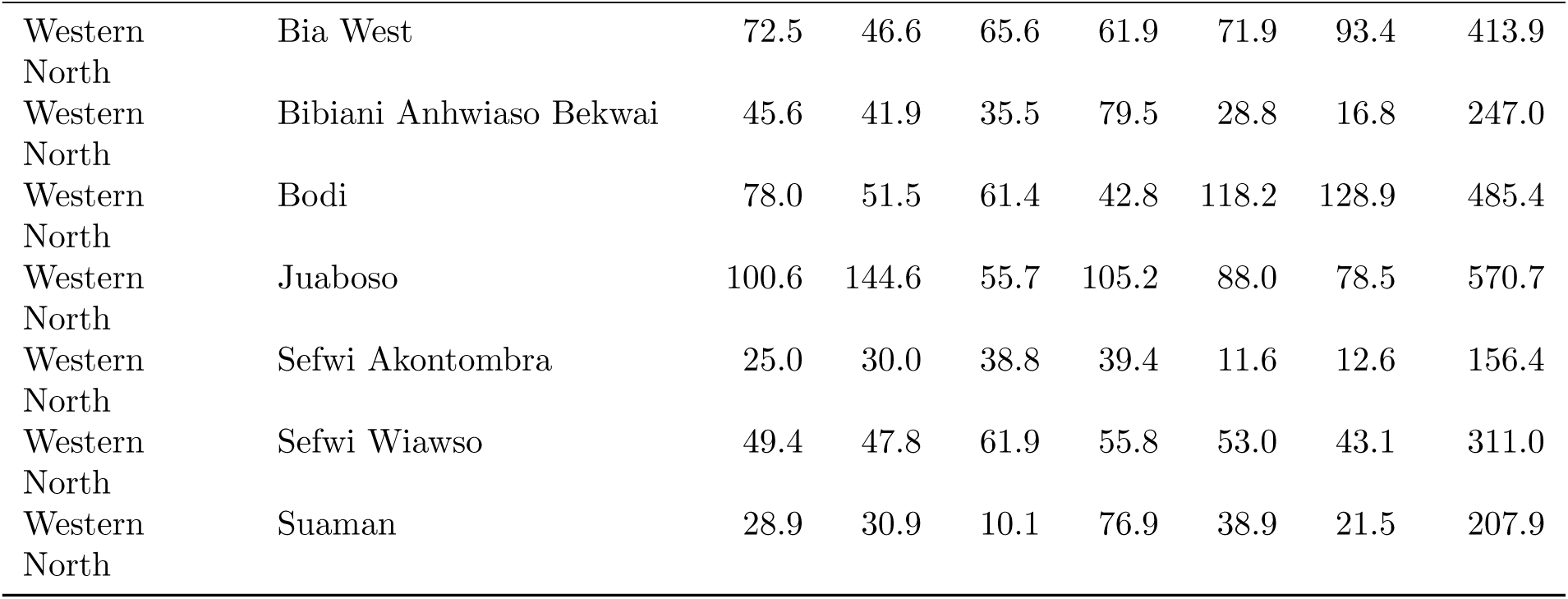
Observed annual snakebite incidence per 100,000 population by district and region, Ghana, 2020–2025.

**Table A2:**
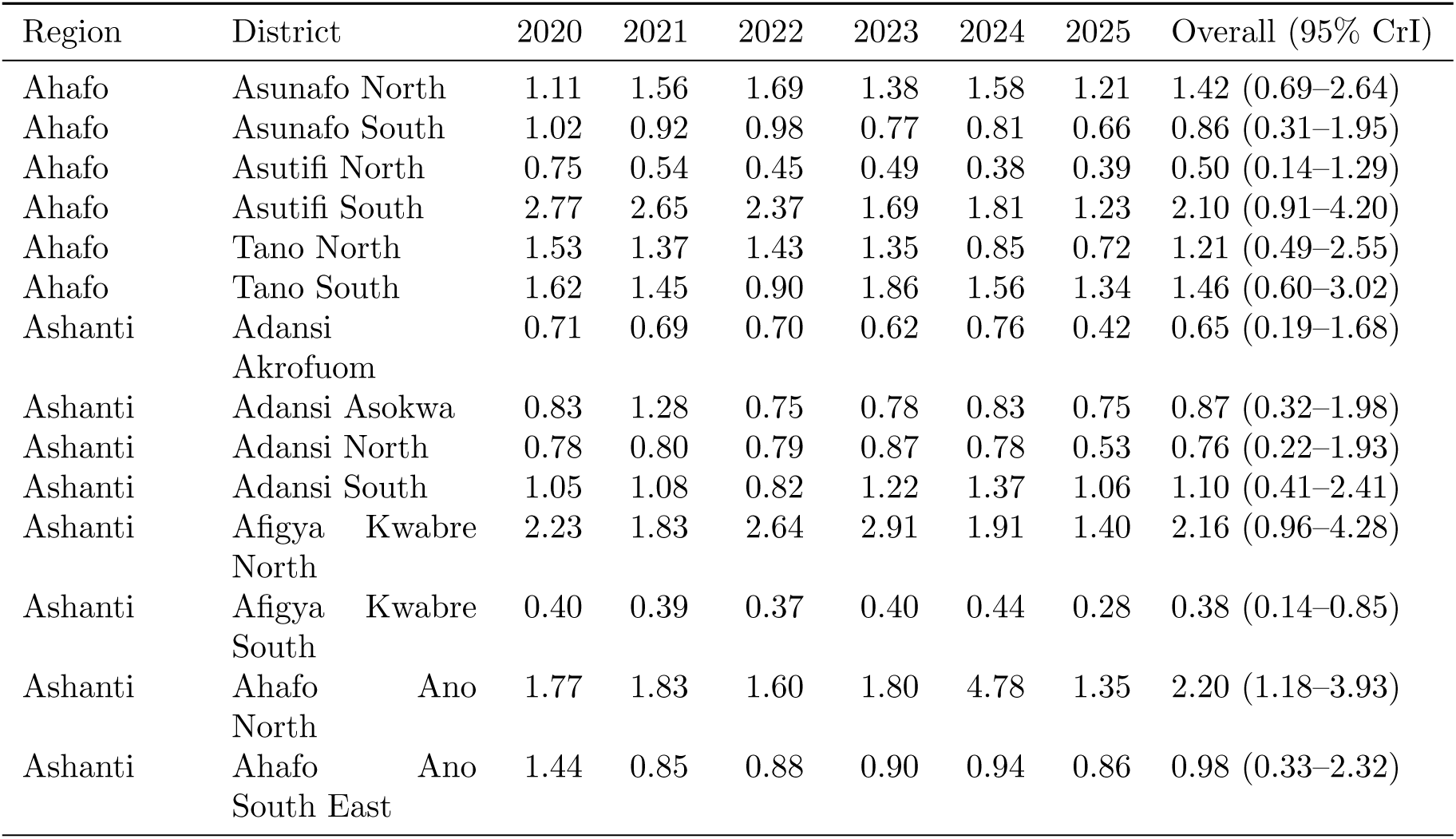
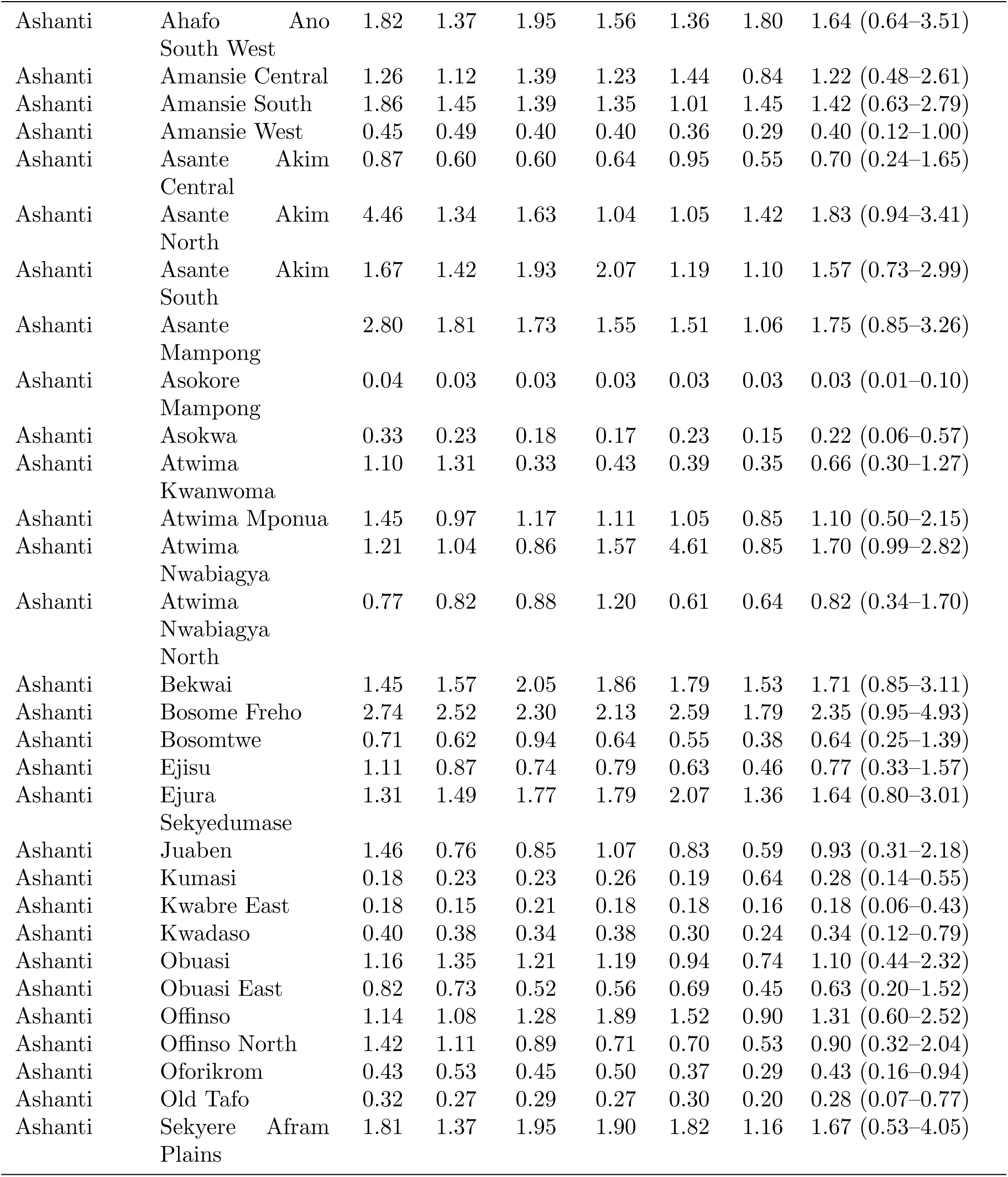
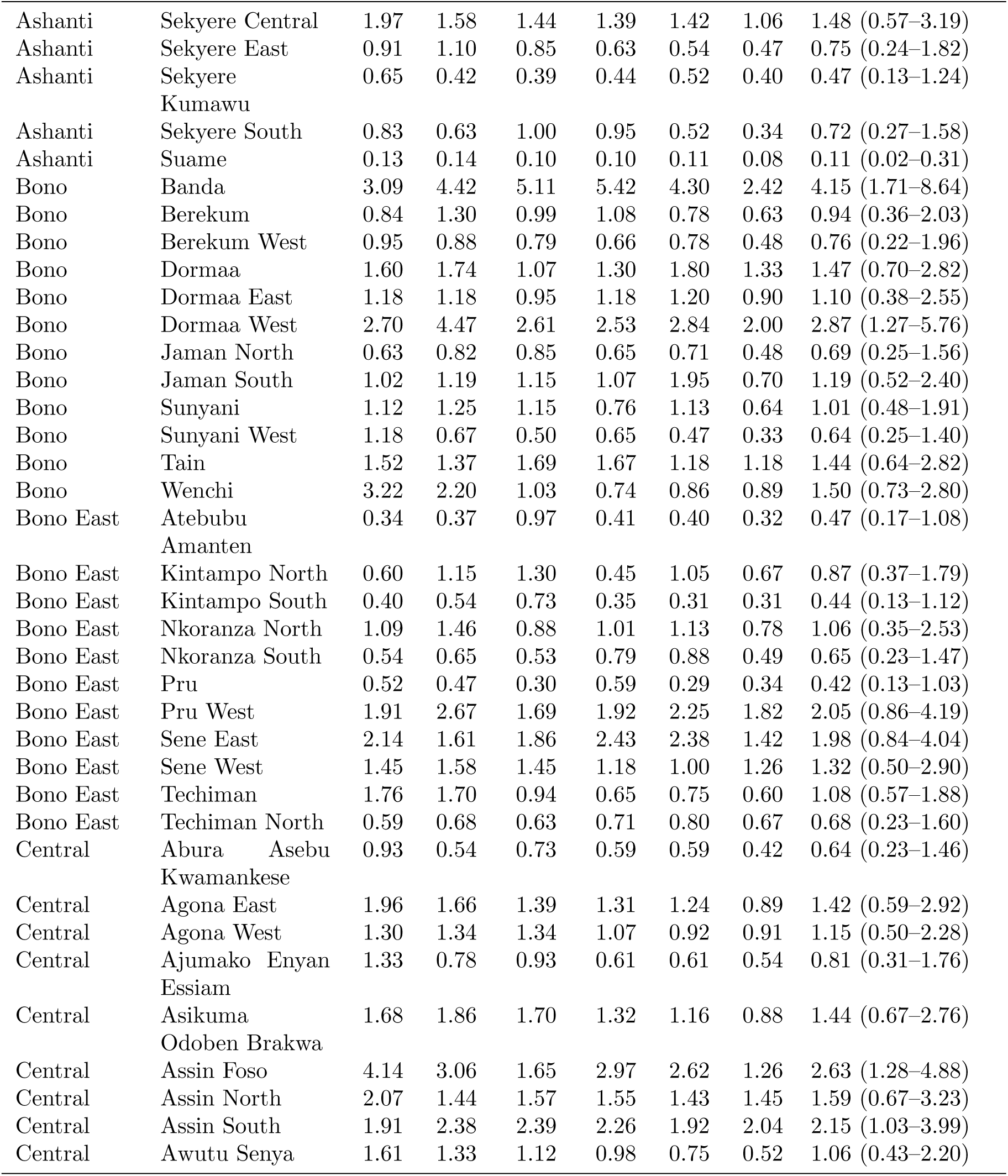
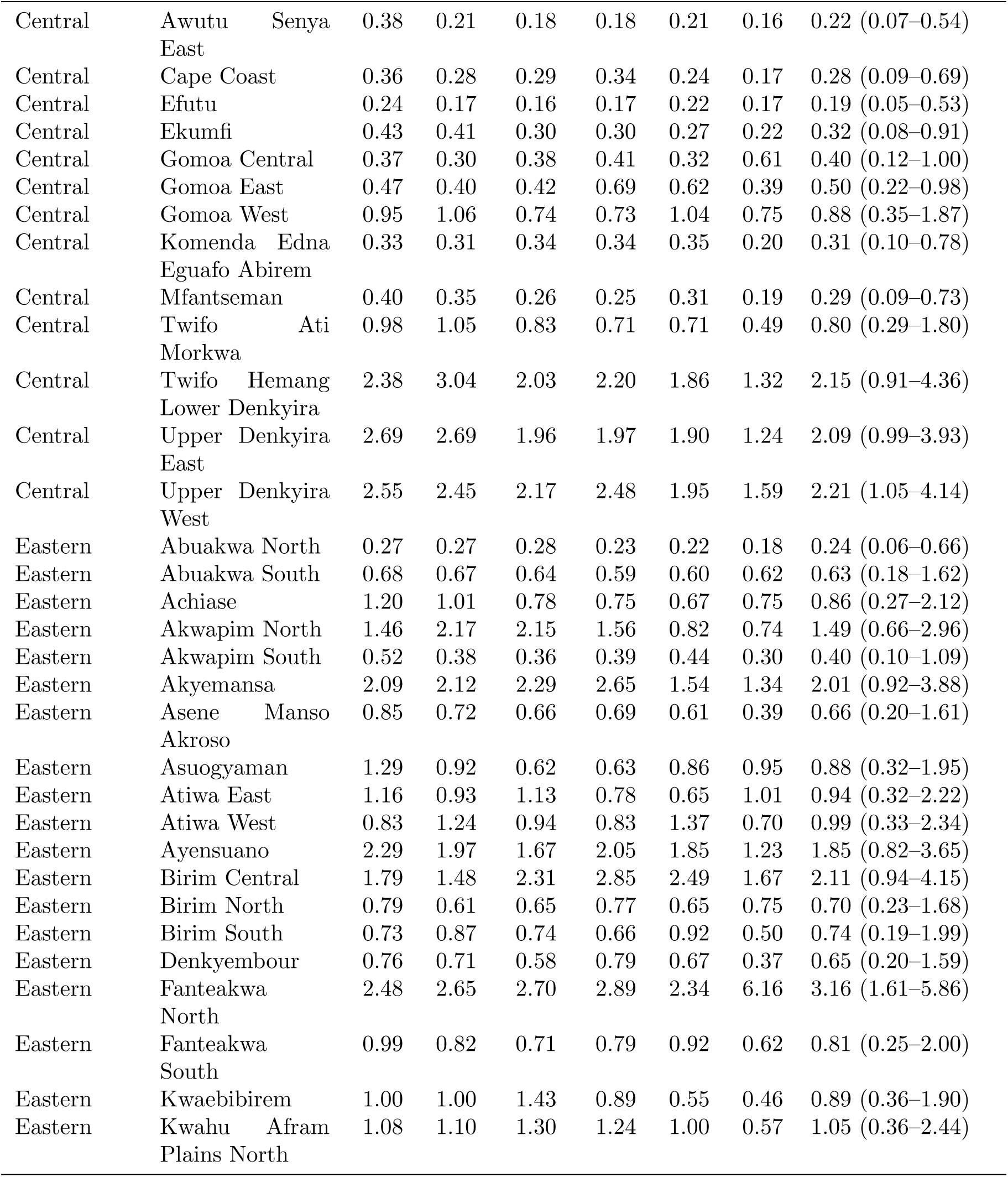
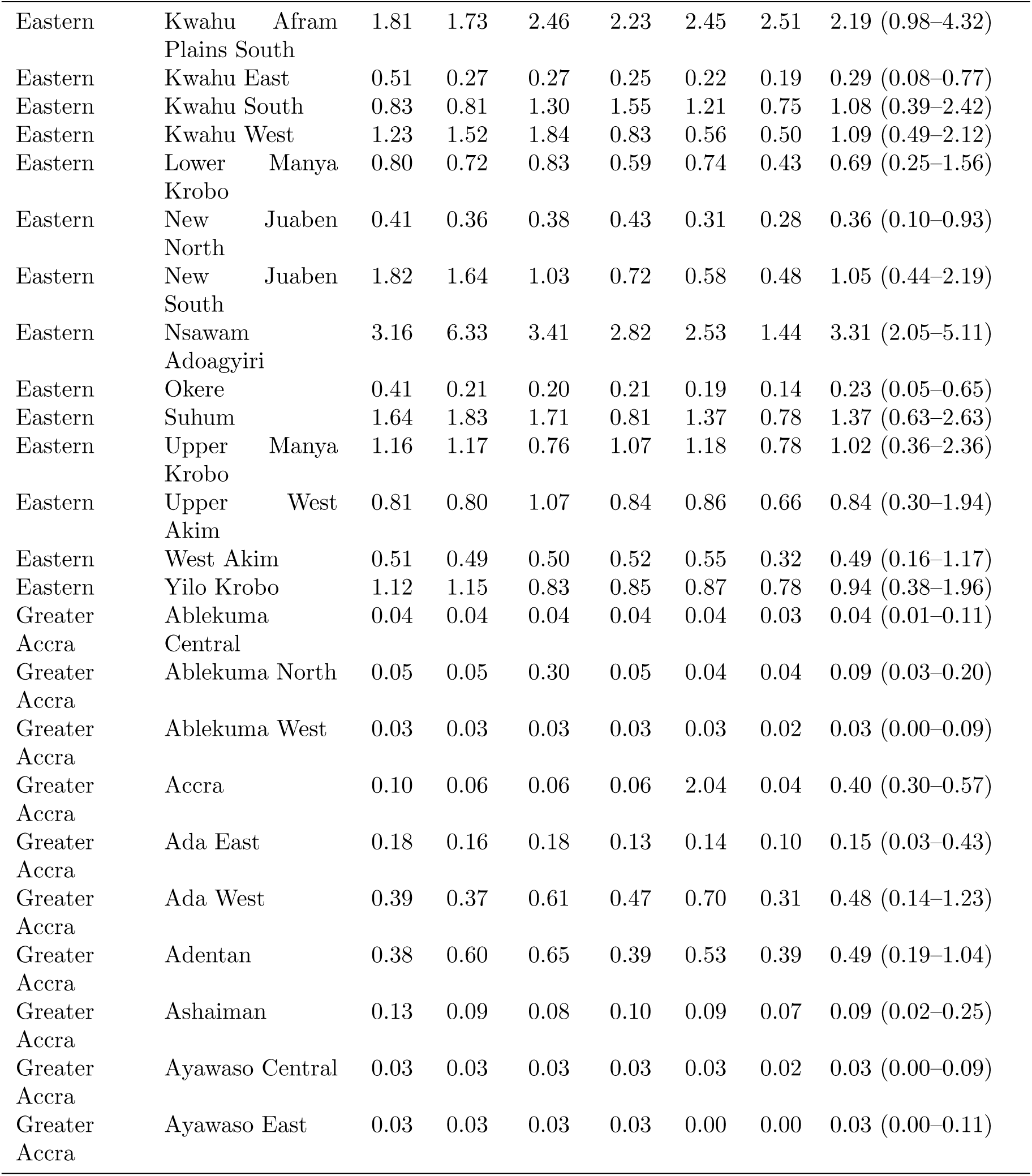
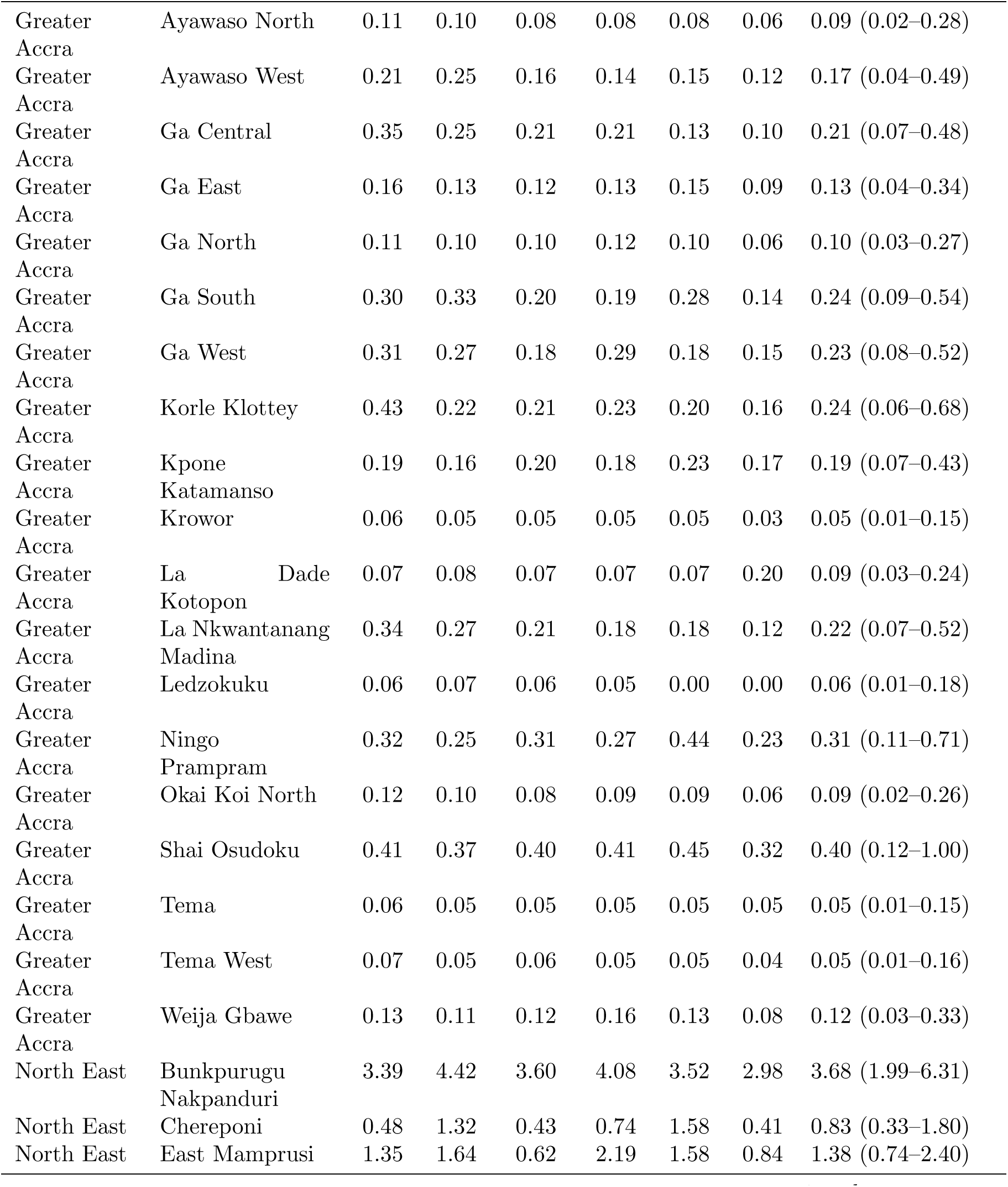
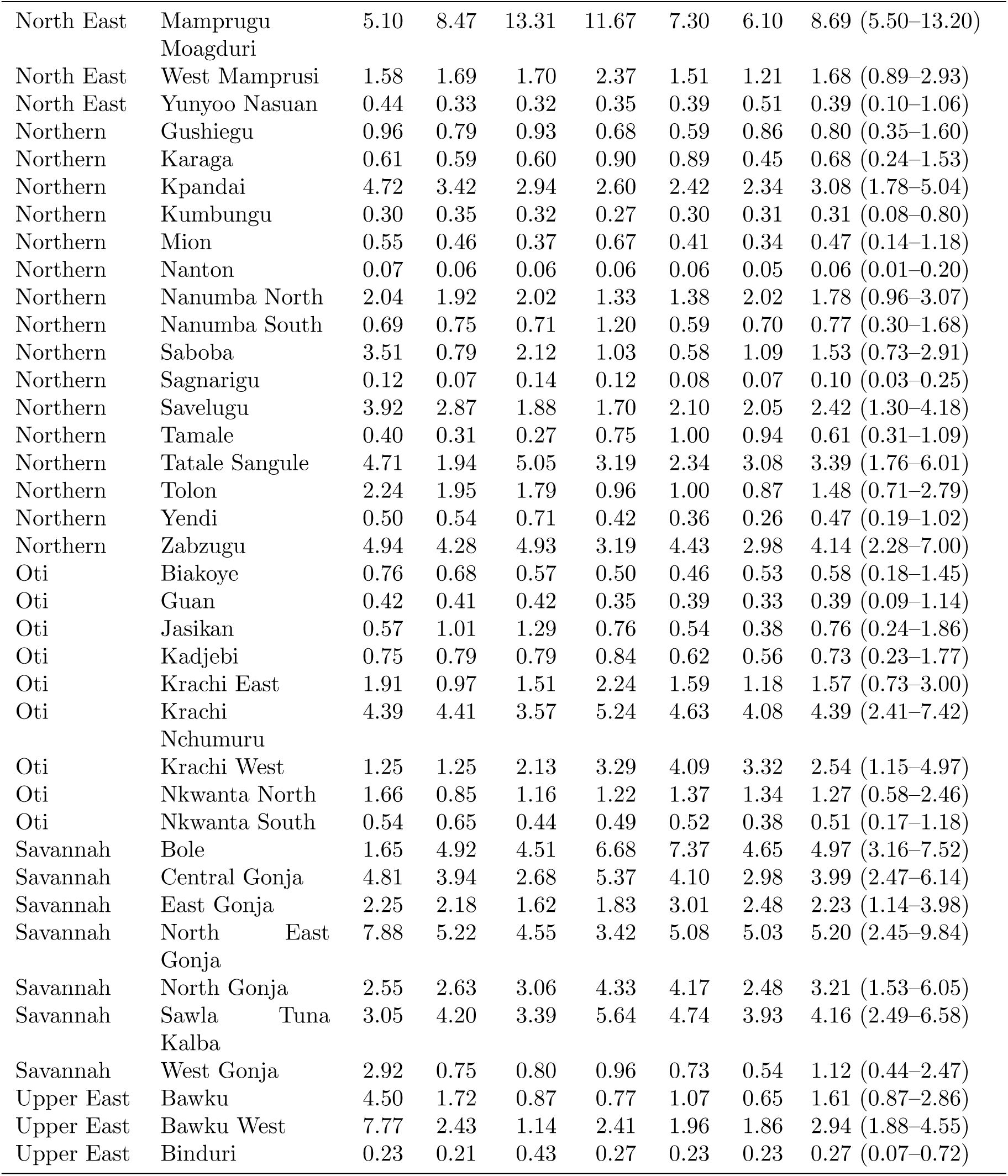
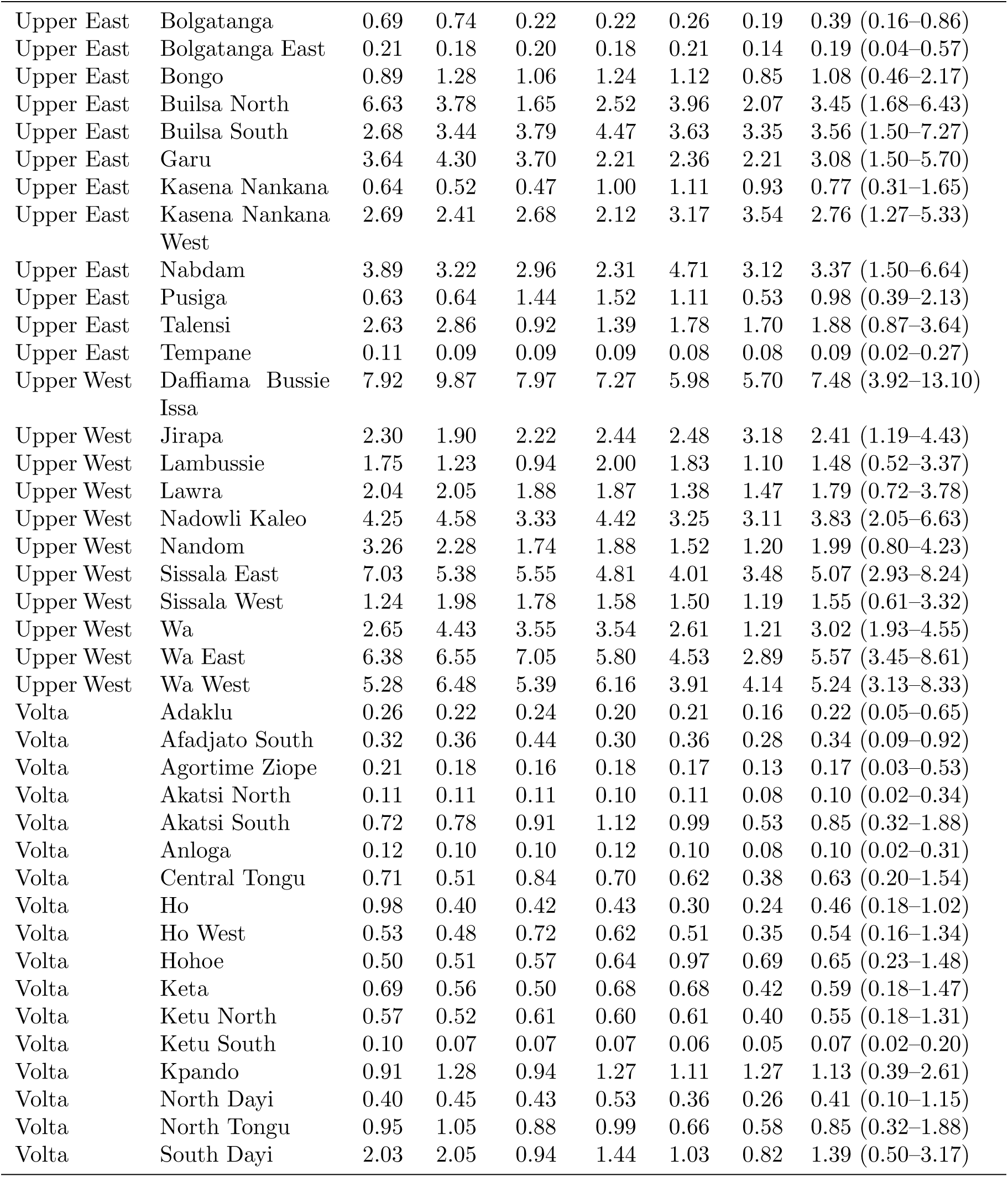
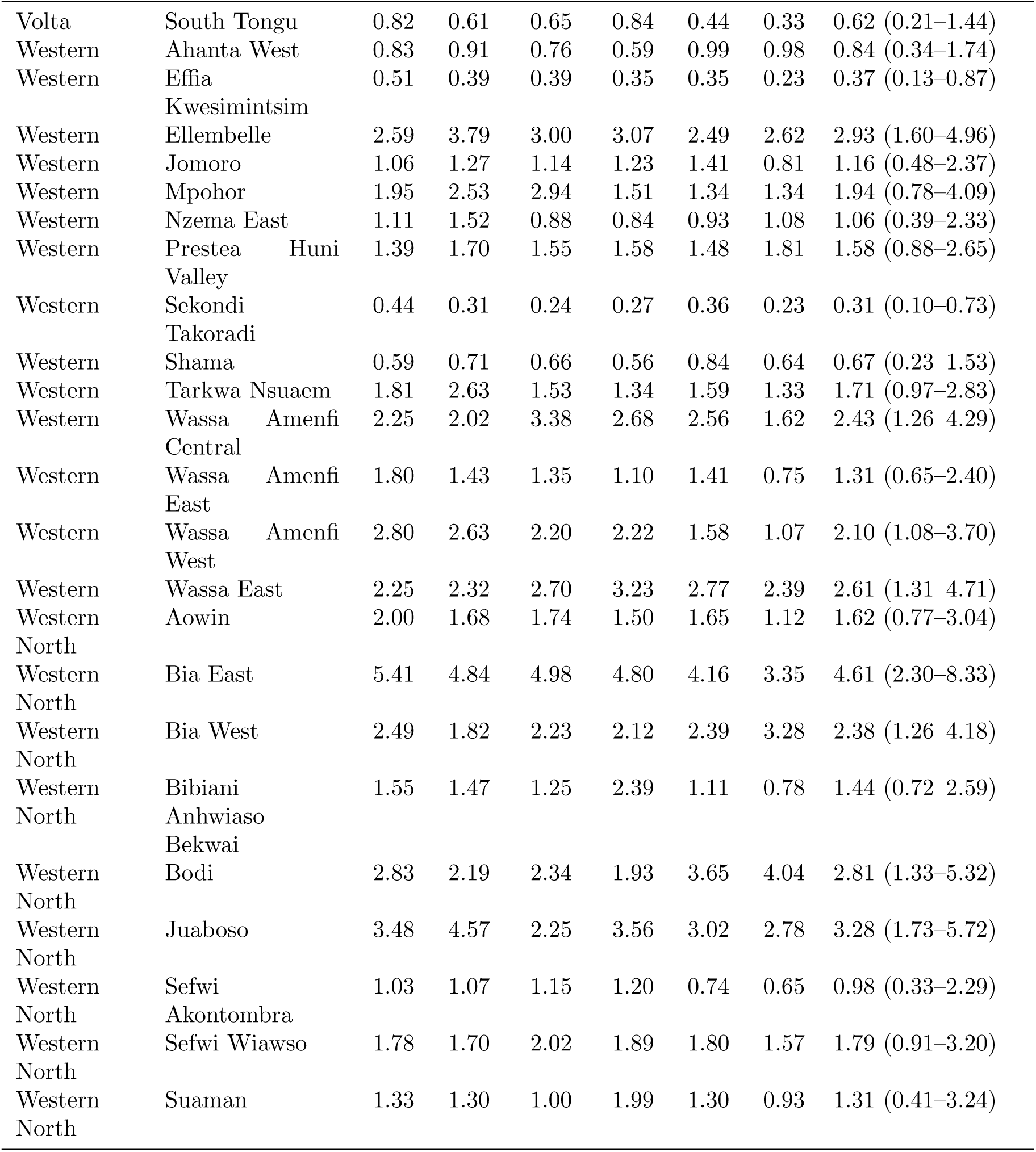
District-level relative risk (RR) of snakebite by year, Ghana, 2020–2025, with the overall (period-averaged) RR and 95% credible interval (CrI), from the Bayesian spatio-temporal BYM2 model.

**Table A3:**
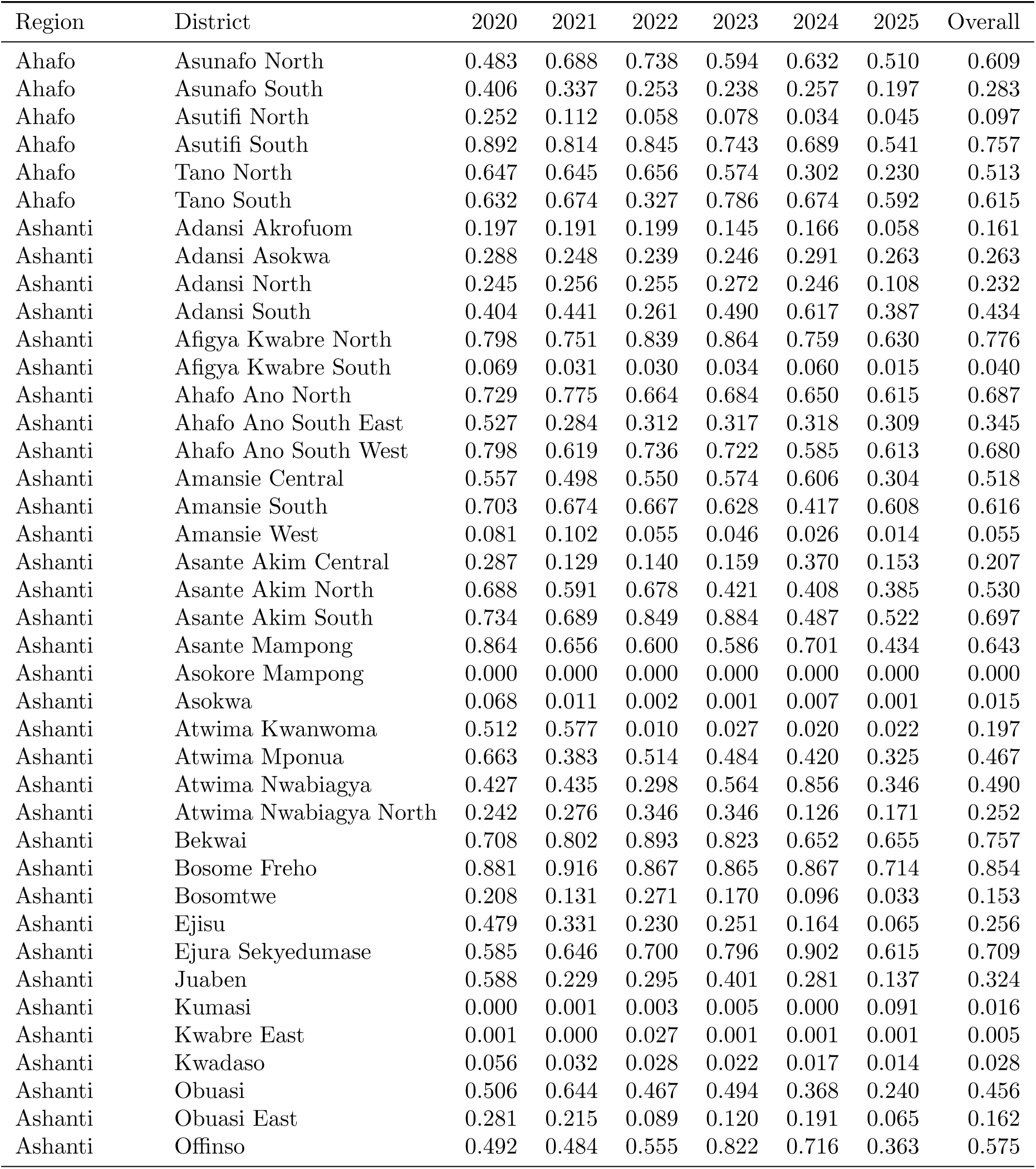
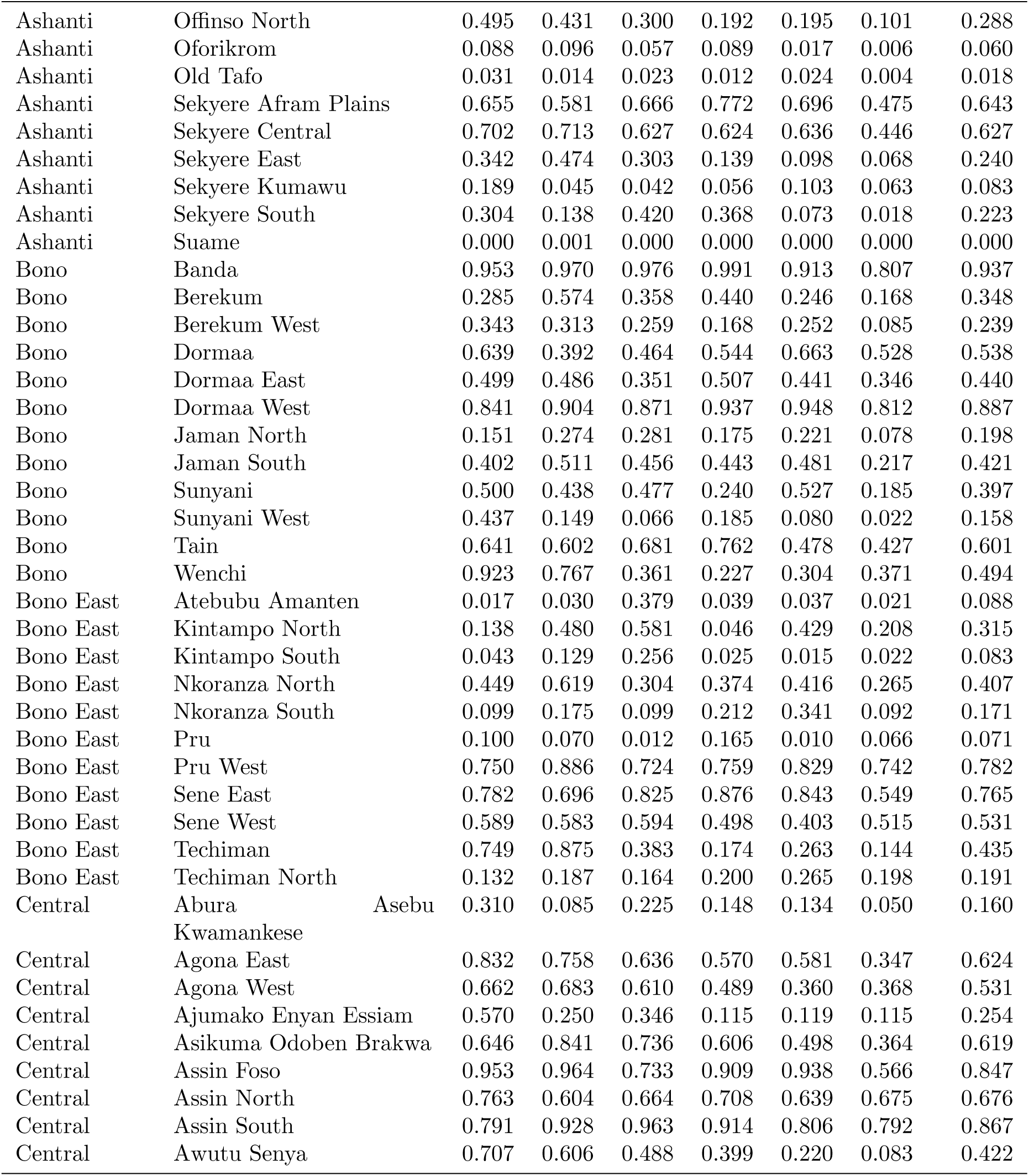
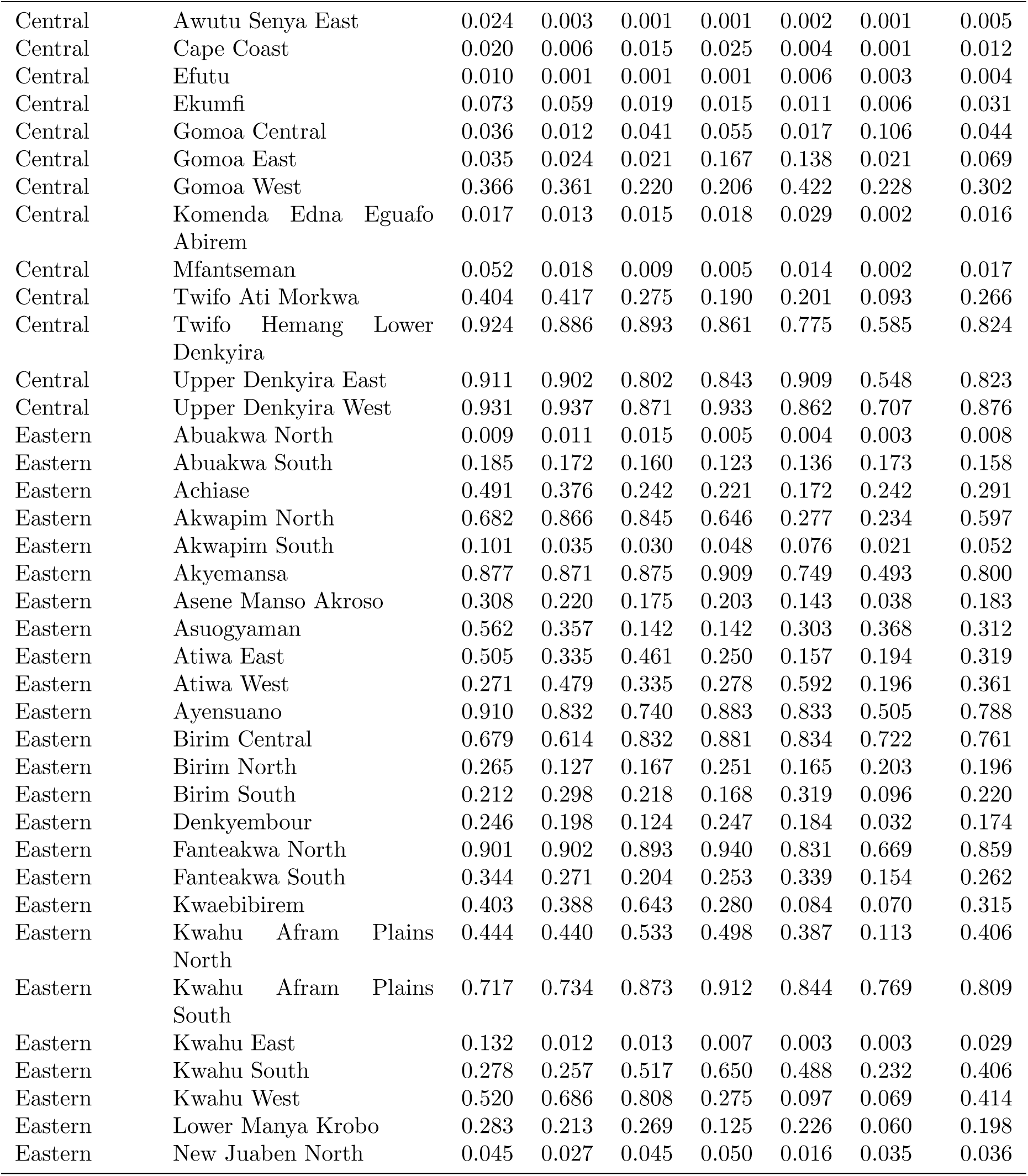
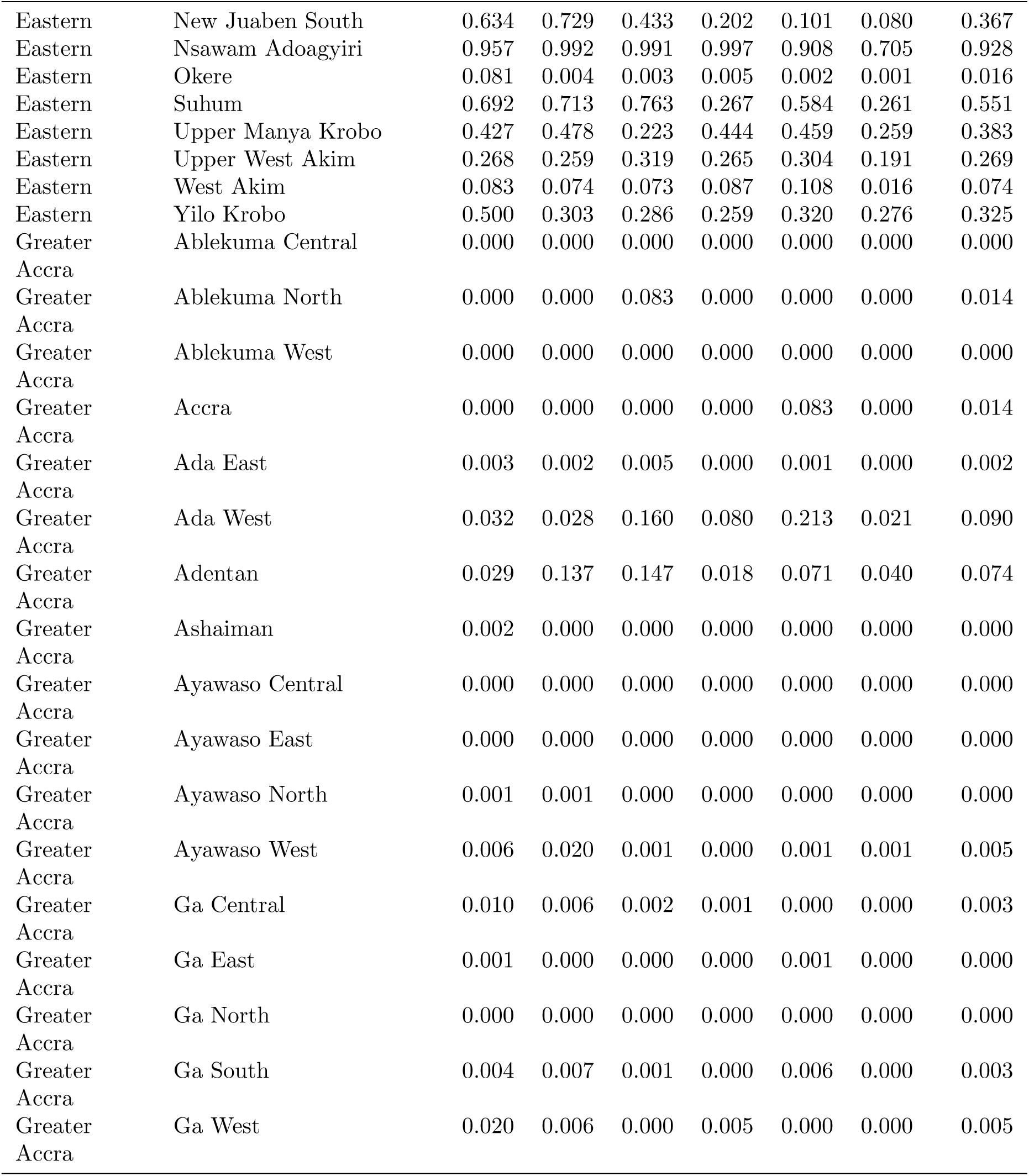
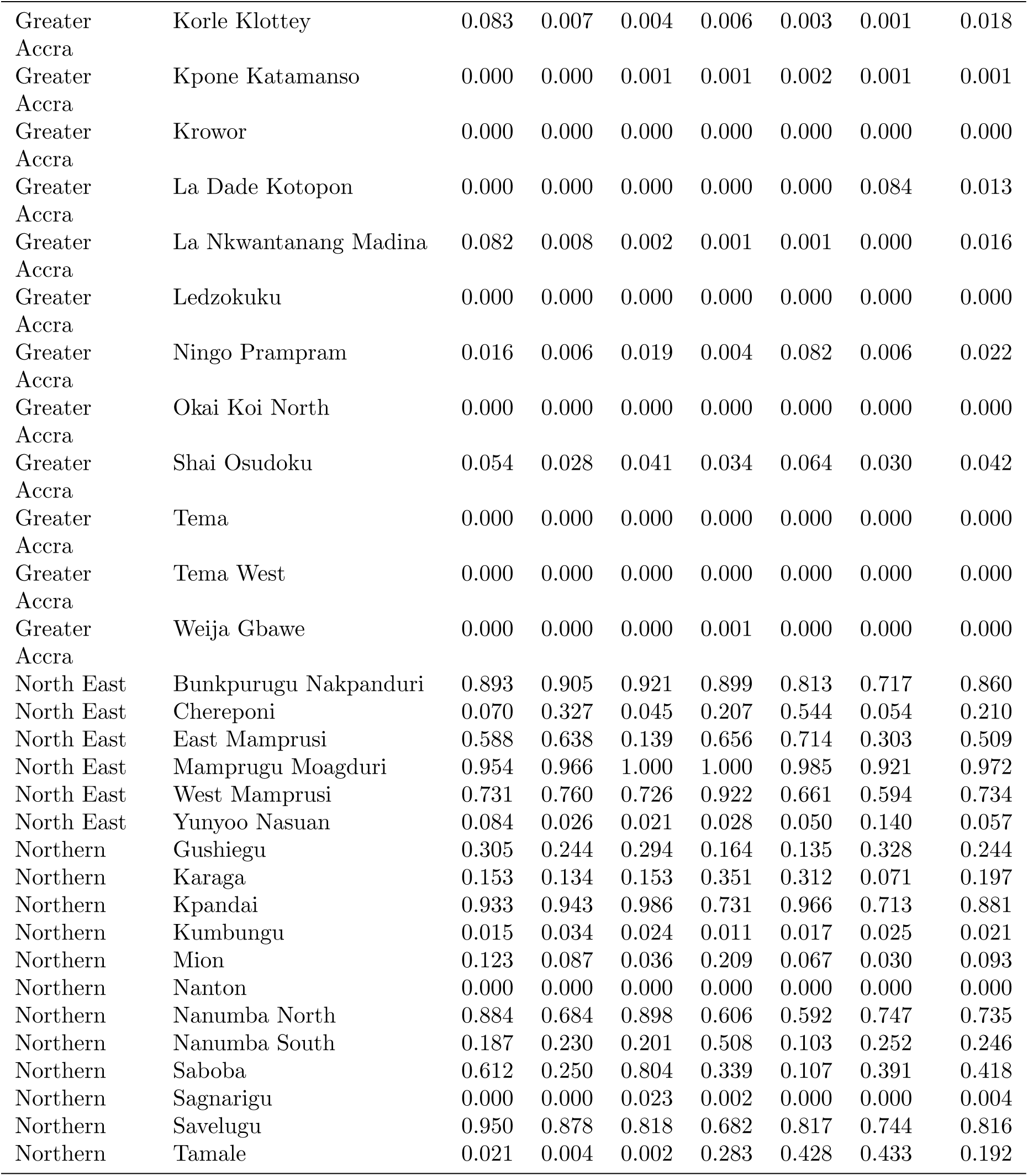
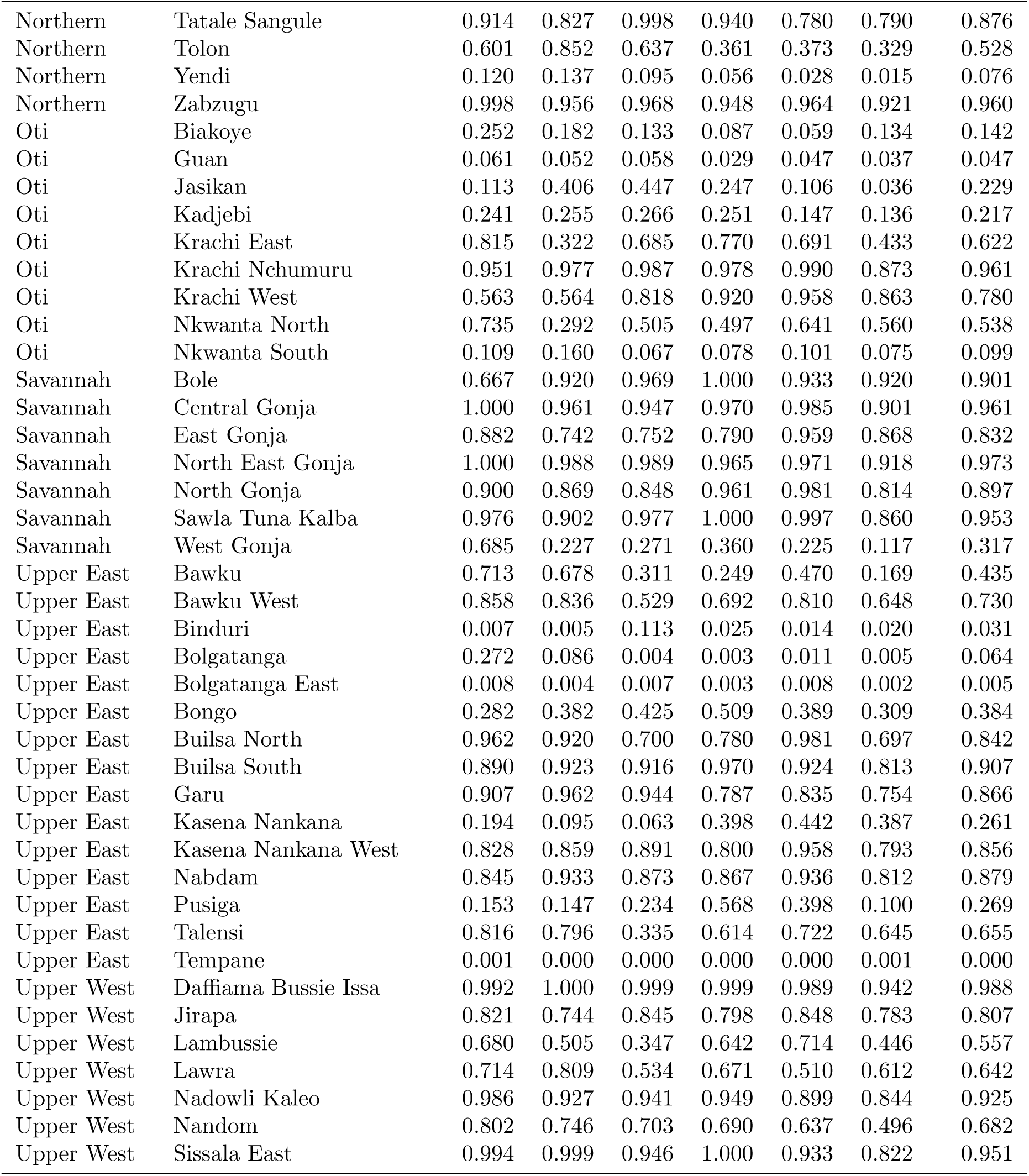
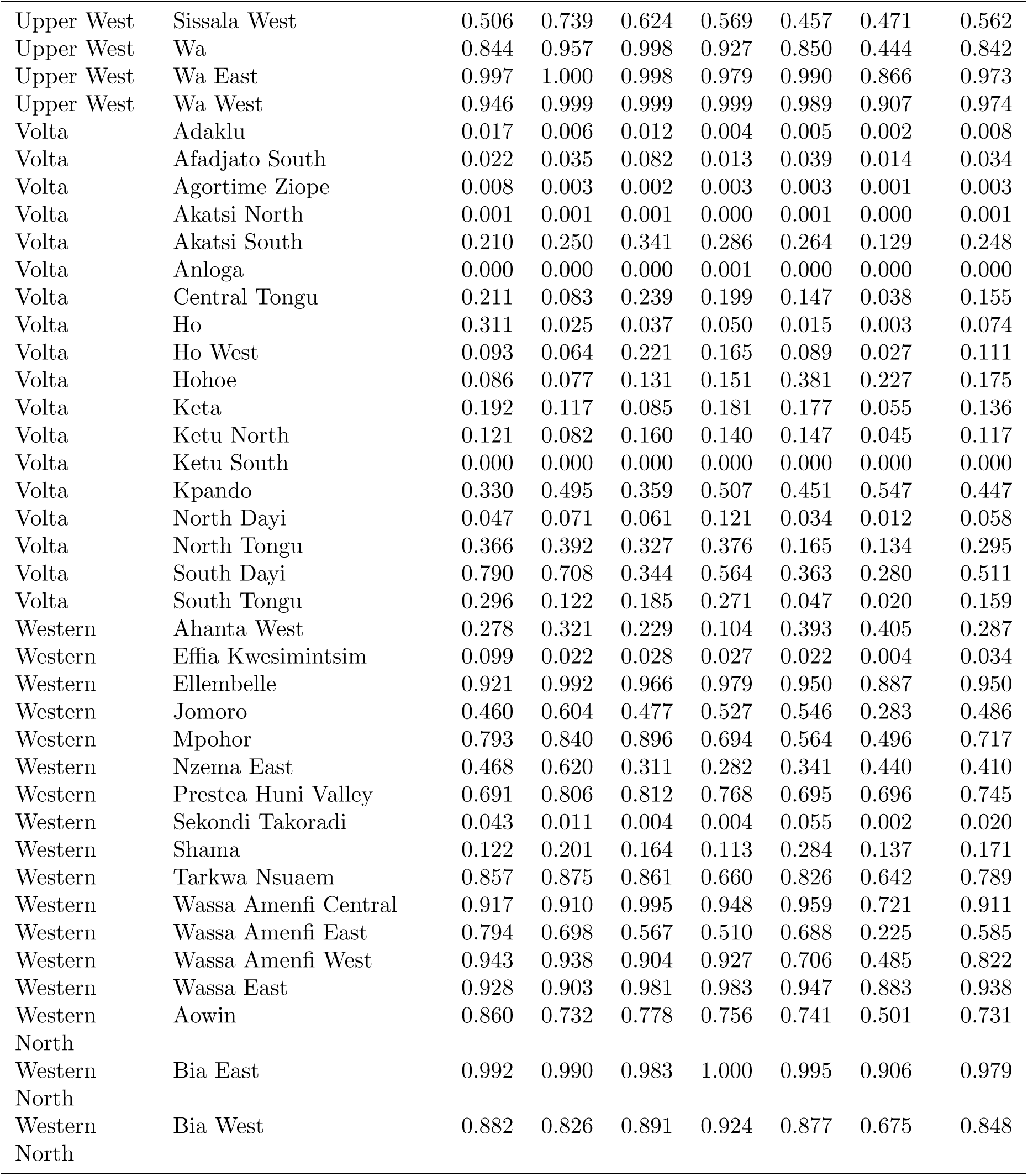
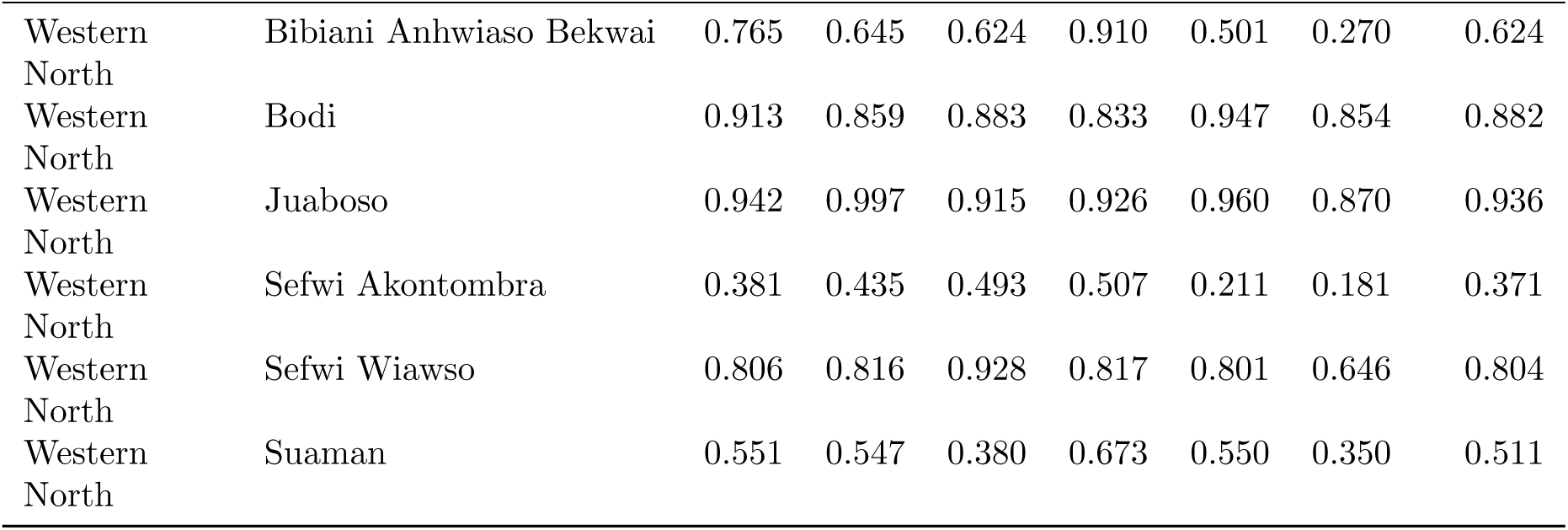
Probability of exceeding the high-incidence threshold by district and region, Ghana, 2020–2025.

**Table A4:**
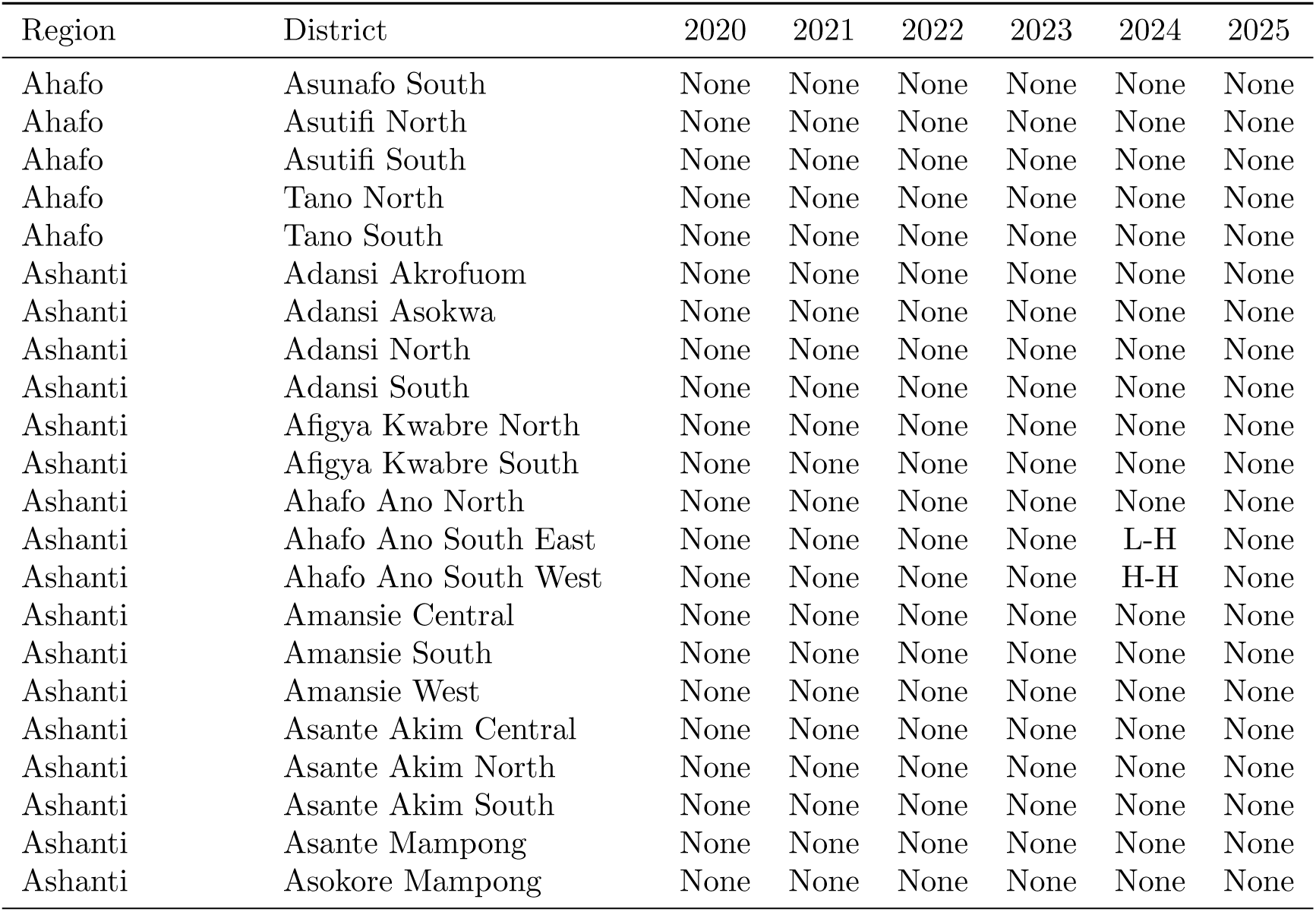
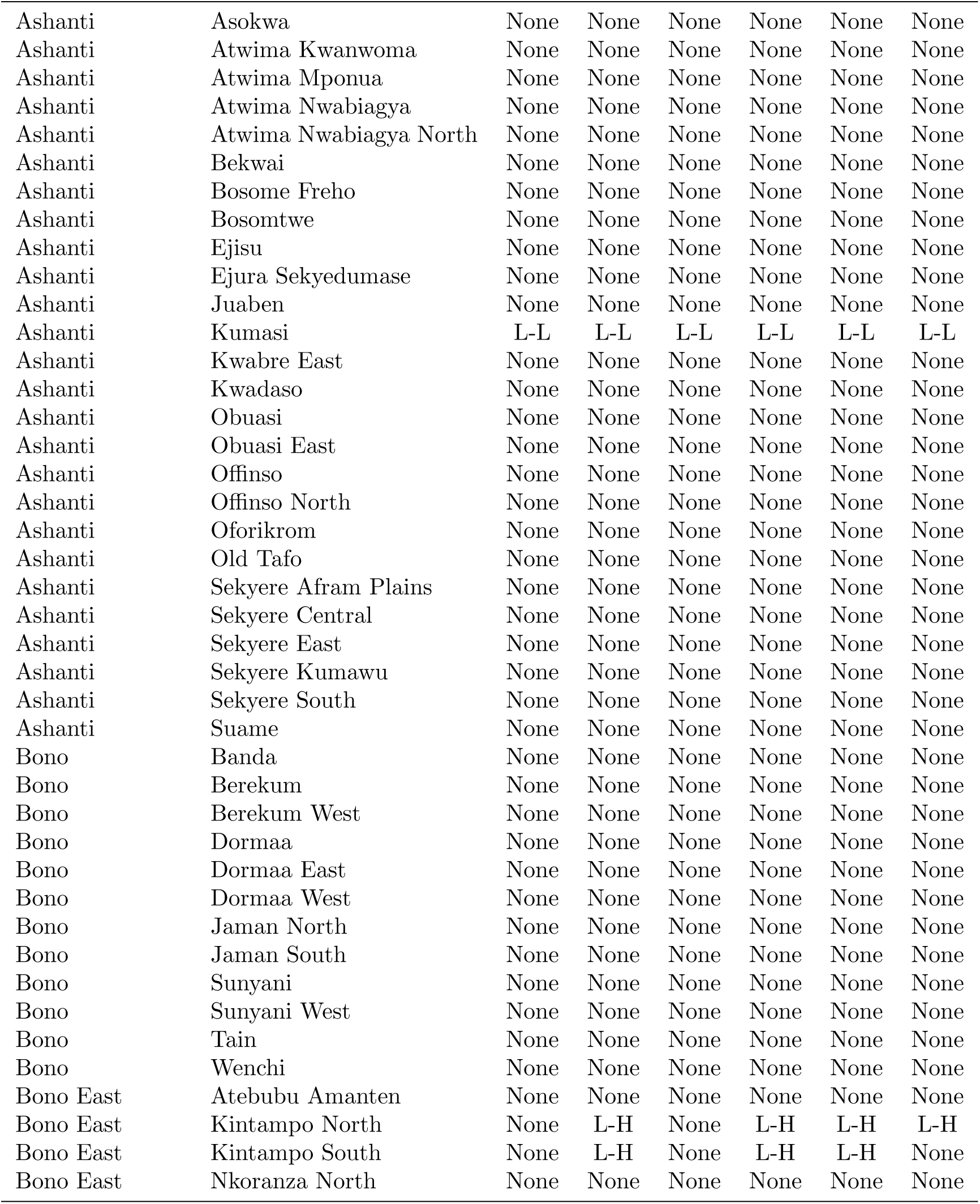
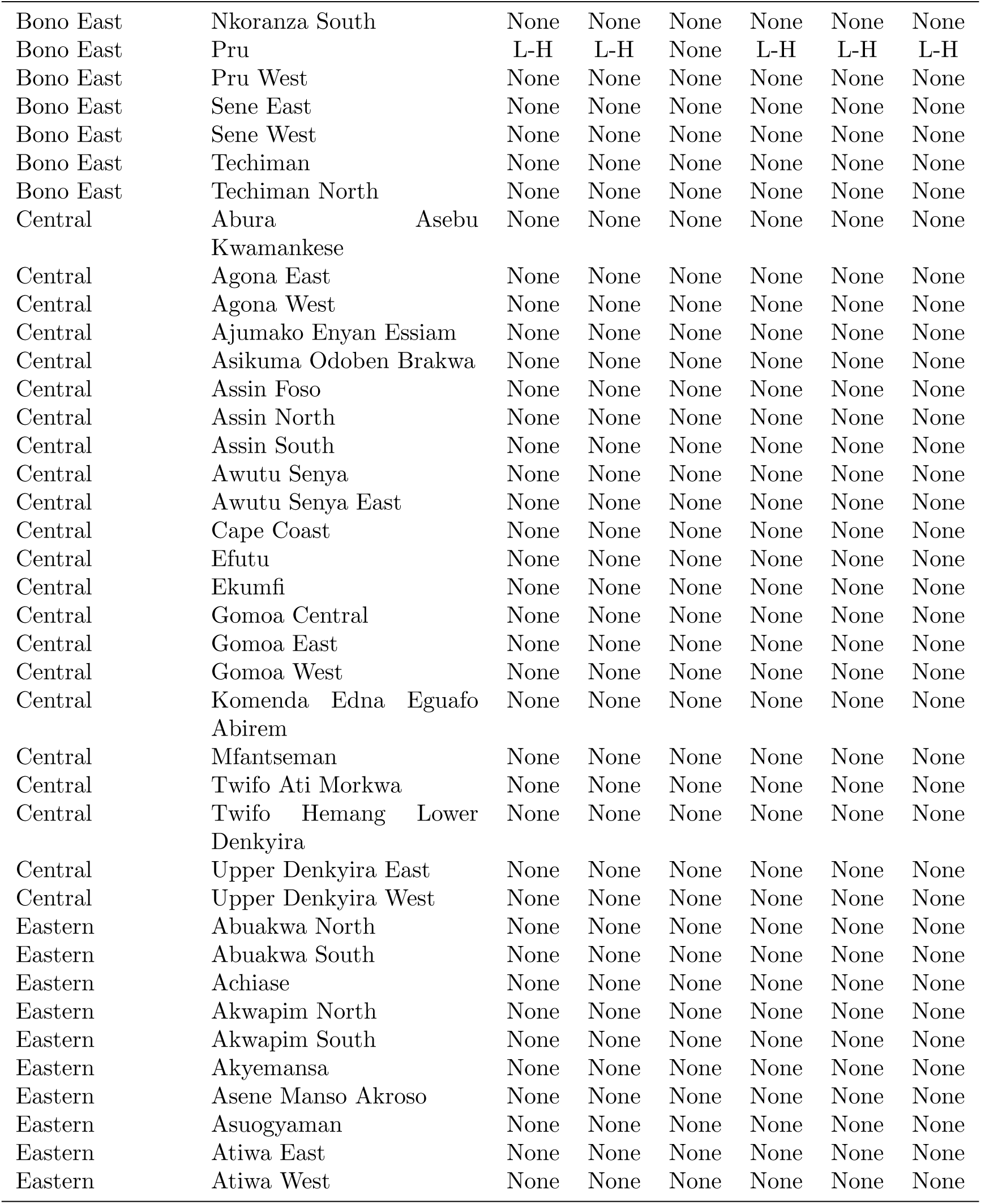
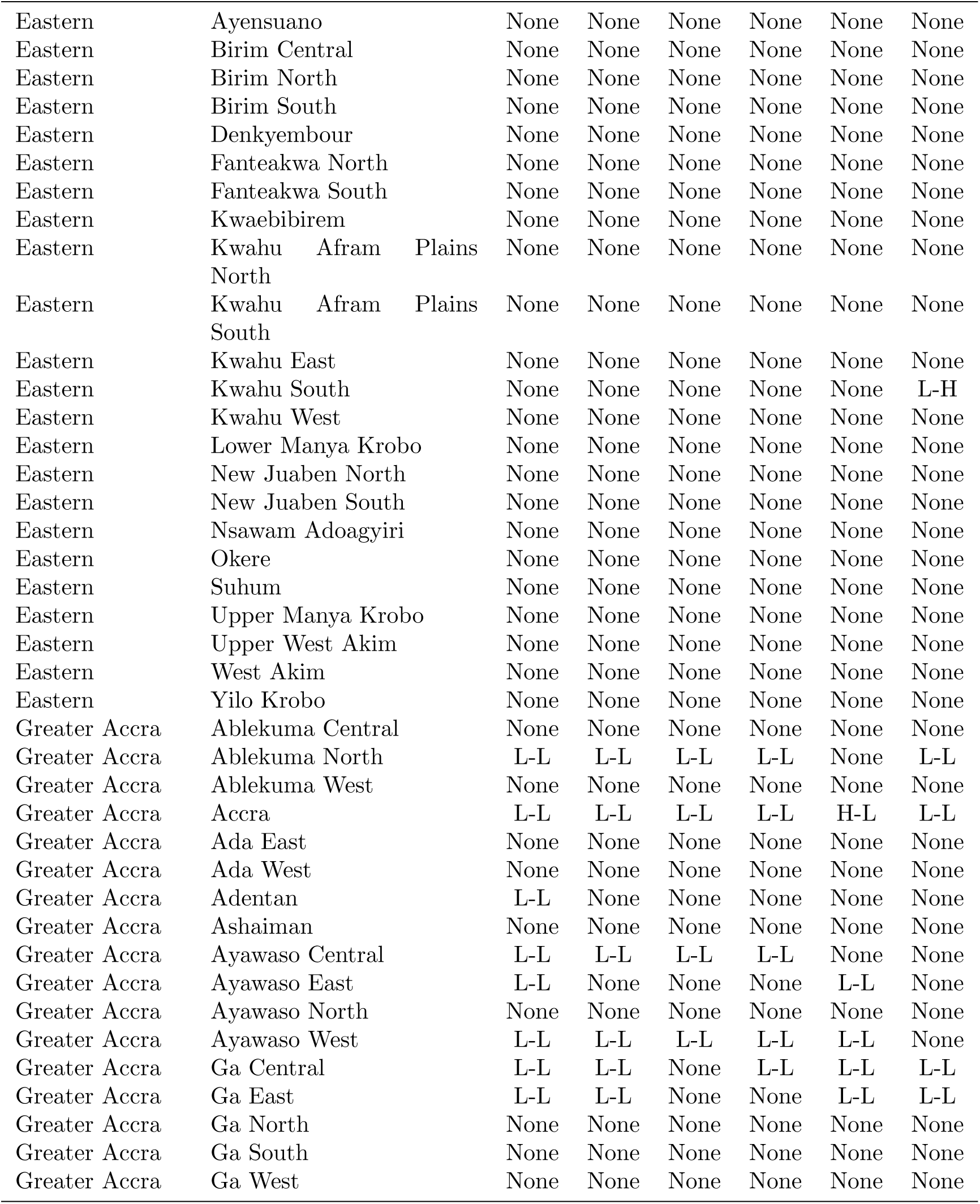
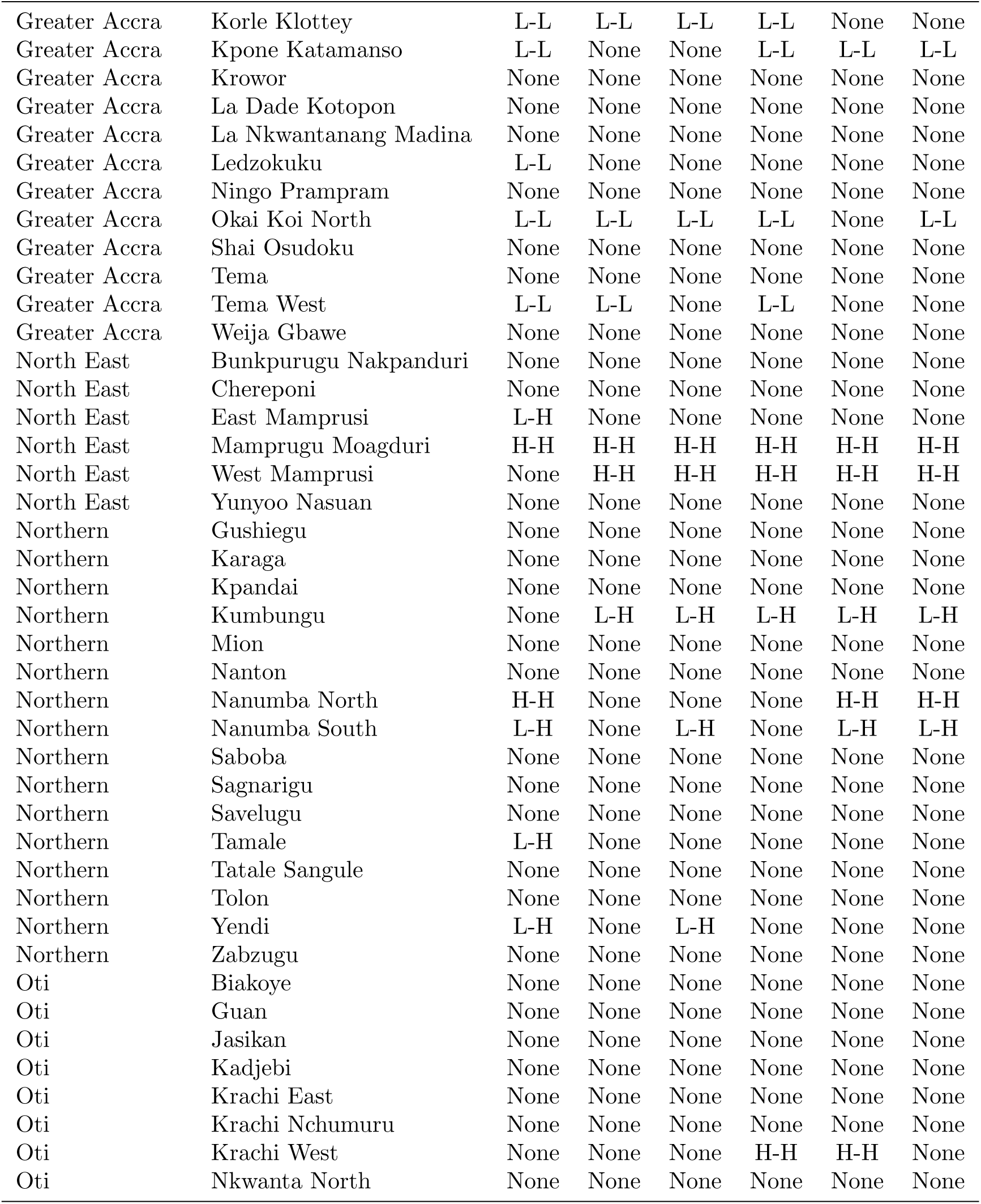
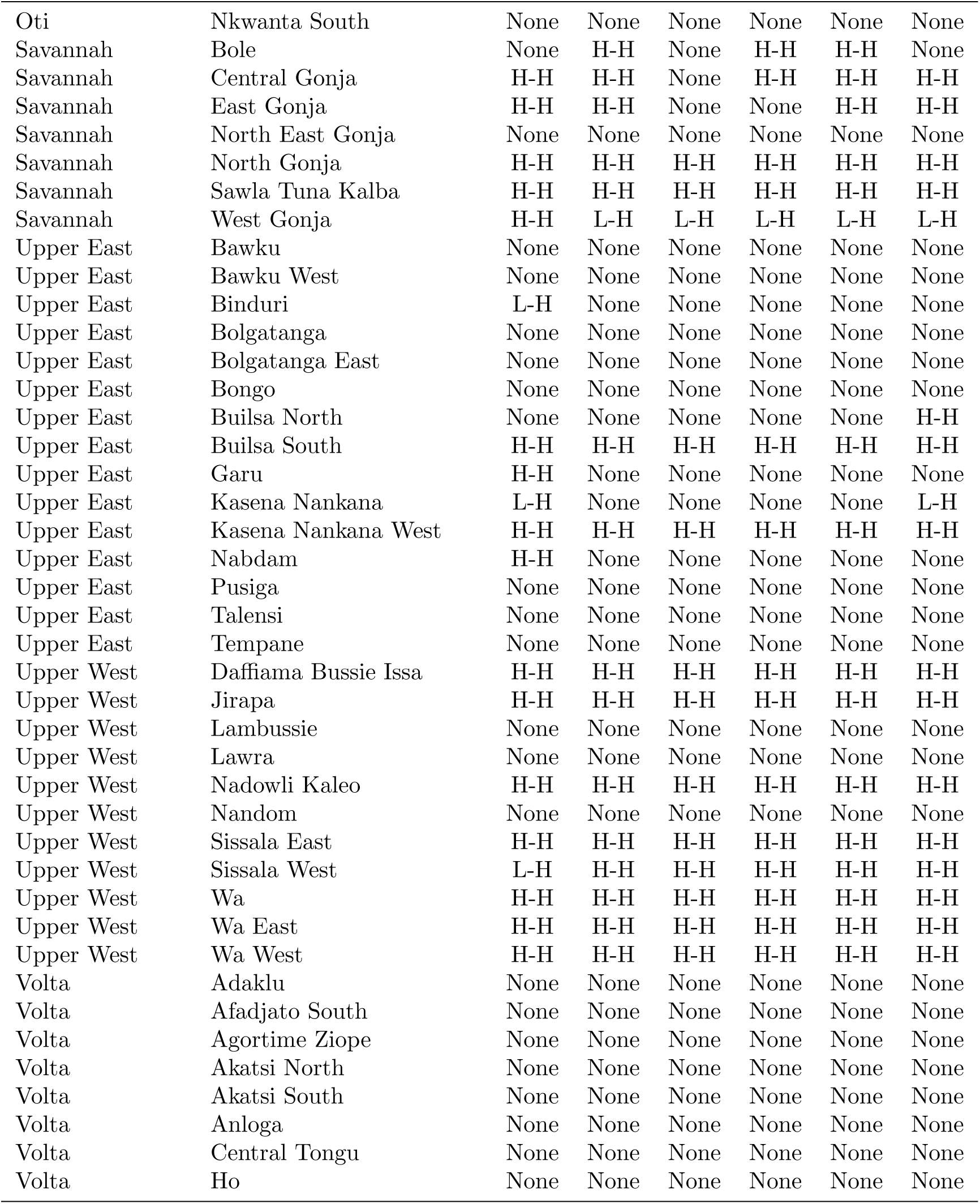
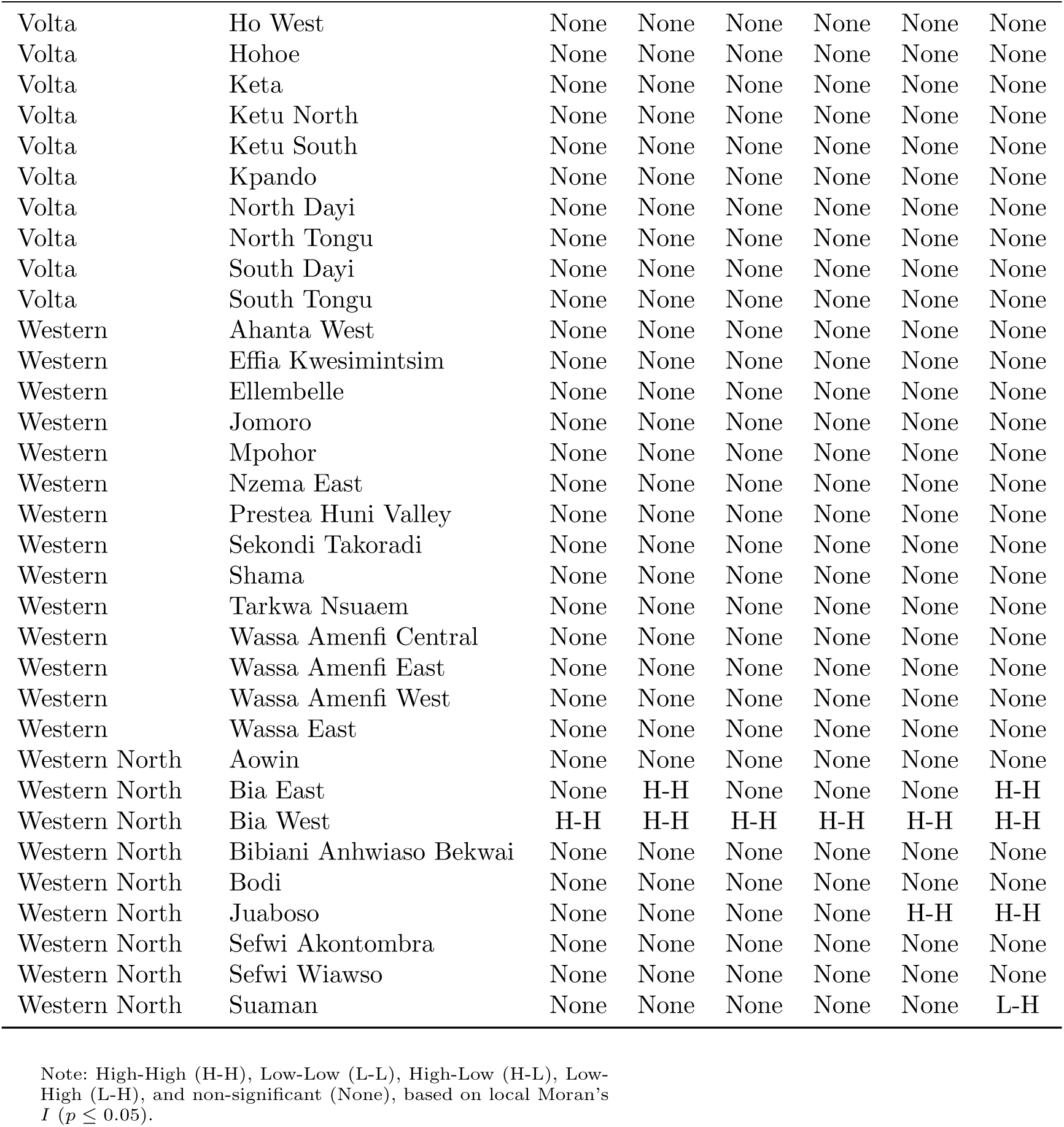
Local Indicators of Spatial Association cluster classification of snakebite relative risk by district and region, Ghana, 2020–2025.

## References

[1] Munshi, H., Gajbhiye, R.K.: Strengthening global snakebite data for who’s goal for 2030. The Lancet 403(10430), 907–908 (2024)

[2] GBD 2019 Snakebite Envenomation Collaborators, Charlson, F.J., Diaz, V., Dirac, M.A., Fliatouris, M., Glenn, S.D., Lalloo, D.G., Salimi, Y., Welgan, C.A., Harrison, R.A., Hay, S.I., Murray, C.J.L., Naghavi, M.: Global mortality of snakebite envenoming between 1990 and 2019. Nature Communications **13**(1), 6160 (2022) 10.1038/s41467-022-33627-9

[3] N’Krumah, T.A.S.R., Końe, B.V., Koffi, Y.D., Coulibaly, D.I., Tall, A., Końe, S., Toppino, S., Stojkovic, M., Bonfoh, B., Junghanss, T.: Survey on the burden, epidemiological and clinical characteristics of snakebite envenoming in the health demographic surveillance system (hdss) of taabo (southern ĉote d’ivoire). PLOS Neglected Tropical Diseases 19(4), 0012983 (2025)

[4] Habib, A.G., Kuznik, A., Hamza, M., Abdullahi, M.I., Chedi, B.A., Chippaux, J.-P., Warrell, D.A.: Snakebite is under appreciated: appraisal of burden from west africa. PLoS neglected tropical diseases 9(9), 0004088 (2015)

[5] Ammouch, K., Mesmoudi, N., Hammani, N., Galan, J., Moustaghfir, A., Stöcklin, R., Oukkache, N.: Tackling the burden of envenomation in africa: advances, challenges, and strategic priorities for enhanced diagnosis and treatment. Frontiers in Tropical Diseases 6, 1653213 (2025)

[6] Abanga, W.A., Kwabena, A.S., Adodoo, J., Issahaku, G.R., Akowuah, G., Odikro, M.A., Bando, D.A.B., Kenu, E., Kubio, C.: Geospatial and temporal pattern of reported snakebite cases to health facilities in the savannah region, ghana, 2018–2022. Journal of Interventional Epidemiology and Public Health 8(2), 1–13 (2025)

[7] Deikumah, J.P., Biney, R.P., Awoonor-Williams, J.K., Gyakobo, M.K.: Compendium of medically important snakes, venom activity and clinical presentations in ghana. PLOS Neglected Tropical Diseases 17(7), 0011050 (2023)

[8] Ketor, C.E., Benneh, C.K., Mensah, K.B., Sarkodie, E., Mensah, A., Somuah, S.O., Akakpo, S., Buabeng, K.O.: Snakebites and antisnake venom utilization in ghana’s oti region: A 6-year retrospective study. BioMed Research International 2024(1), 6692421 (2024)

[9] Ochoa, C., Pittavino, M., Babo Martins, S., Alcoba, G., Bolon, I., Castañeda, R., Joost, S., Sharma, S.K., Chappuis, F., Ray, N.: Estimating and predicting snakebite risk in the terai region of nepal through a high-resolution geospatial and one health approach. Scientific reports 11(1), 23868 (2021)

[10] Besag, J., York, J., Mollíe, A.: Bayesian image restoration, with two applications in spatial statistics. Annals of the institute of statistical mathematics 43(1), 1–20 (1991)

[11] Riebler, A., Sørbye, S.H., Simpson, D., Rue, H.: An intuitive bayesian spatial model for disease mapping that accounts for scaling. Statistical methods in medical research 25(4), 1145–1165 (2016)

[12] [12] Simpson, D., Rue, H., Riebler, A., Martins, T.G., Sørbye, S.H.: Penalising model component complexity: A principled, practical approach to constructing priors (2017)

[13] Knorr-Held, L.: Bayesian modelling of inseparable space-time variation in disease risk. Statistics in medicine 19(17-18), 2555–2567 (2000)

[14] Satorra, P., Tebé, C.: Bayesian spatio-temporal analysis of the covid-19 pandemic in catalonia. Scientific Reports 14(1), 4220 (2024)

[15] Moran, P.A.: Notes on continuous stochastic phenomena. Biometrika 37(1/2), 17–23 (1950)

[16] Anselin, L.: Local indicators of spatial association—lisa. Geographical analysis 27(2), 93–115 (1995)

[17] Yousefi, M., Yousefkhani, S.H., Grünig, M., Kafash, A., Rajabizadeh, M., Pouyani, E.R.: Identifying high snakebite risk area under climate change for community education and antivenom distribution. Scientific Reports 13(1), 8191 (2023)

[18] Rangel-Camacho, R., Ýañez-Arenas, C., Chippaux, J., Martın, G.: Socioeconomic and ecological drivers of snakebite incidence in mexico: A spatial analysis of risk factors. PLOS Neglected Tropical Diseases 19(10), 0013582 (2025)

[19] Collinson, S., Lamb, T., Cardoso, I.A., Diggle, P.J., Lalloo, D.G.: A systematic review of variables associated with snakebite risk in spatial and temporal analyses. Transactions of the Royal Society of Tropical Medicine and Hygiene 119(9), 1084–1099 (2025)

[20] Bravo-Vega, C., Santos-Vega, M., Cordovez, J.M.: Disentangling snakebite dynamics in colombia: how does rainfall and temperature drive snakebite temporal patterns? PLoS Neglected Tropical Diseases 16(3), 0010270 (2022)

[21] Ghana Statistical Service: Population and housing census: General report. Technical report, Ghana Statistical Service (GSS), Accra, Ghana (2021)

[22] GADM: GADM Database of Global Administrative Areas, Version 4.1. https://gadm.org. Accessed: July 12, 2026(2022)

[23] WorldPop, Bondarenko, M., Kerr, D., Sorichetta, A., Tatem, A.J.: Census/Projection-Disaggregated Gridded Population Datasets, Adjusted to Match the Corresponding UNPD 2020 Estimates, for 183 Countries in 2020 Using Built-Settlement Growth Model (BSGM) Outputs. University of Southampton. Constrained Individual Countries dataset, 1 km resolution (2020).

[24] Sparks, A.H.: nasapower: A NASA POWER global meteorology, surface solar energy and climatology data client for R. Journal of Open Source Software 3(30), 1035 (2018) 10.21105/joss.01035

[25] Gorelick, N., Hancher, M., Dixon, M., Ilyushchenko, S., Thau, D., Moore, R.: Google Earth Engine: Planetary-scale geospatial analysis for everyone. Remote Sensing of Environment 202, 18–27 (2017) 10.1016/j.rse.2017.06.031

[26] R Core Team: R: A Language and Environment for Statistical Computing. R Foundation for Statistical Computing, Vienna, Austria (2025). R Foundation for Statistical Computing. https://www.R-project.org/

[27] Rue, H., Martino, S., Chopin, N.: Approximate Bayesian inference for latent Gaussian models by using integrated nested Laplace approximations. Journal of the Royal Statistical Society: Series B (Statistical Methodology) 71(2), 319–392 (2009) 10.1111/j.1467-9868.2008.00700.x

[28] Nori, J., et al.: Climate change-related distributional range shifts of venomous snakes: A predictive modelling study of effects on public health and biodiversity. The Lancet Planetary Health 8(3), 163–171 (2024) 10.1016/S2542-5196(24)00005-6

[29] World Health Organization: Snakebite and Climate Change: A Call for Urgent Action to Future-Proof a Neglected Tropical Disease. https://www.who.int/news/item/12-01-2024-snakebite-and-climate-change---a-call-for-urgent-action-to-future-proof-a-neglected-tropica Departmental update. Accessed: 2026-07-05 (2024)

[30] Jesus, F.M., et al.: Environmental drivers of tropical forest snake phenology: Insights from citizen science. Ecology and Evolution 13, 10305 (2023) 10.1002/ece3.10305

[31] Landry, M., D’Souza, R., Moss, S., Chang, H.H., Ebelt, S., Wilson, L., Scovronick, N.: The association between ambient temperature and snakebite in georgia, usa: a case-crossover study. GeoHealth 7(7), 2022–000781 (2023)

[32] Hanback, S., Slattery, A., McGwin, G., Arnold, J.: Association of daily high temperatures with increased snake envenomations: A case-crossover study. Toxicon 201, 54–58 (2021)

[33] Martın, G., Erinjery, J.J., Ediriweera, D., Goldstein, E., Somaweera, R., Silva, H.J., Lalloo, D.G., Iwamura, T., Murray, K.A.: Effects of global change on snakebite envenoming incidence up to 2050: a modelling assessment. The Lancet Planetary Health 8(8), 533–544 (2024)

[34] Lillywhite, H.B.: How Snakes Work: Structure, Function and Behavior of the World’s Snakes. Oxford University Press, ??? (2014)

[35] Dossou, A.J., Fandohan, A.B., Omara, T., Chippaux, J.-P.: Comprehensive review of epidemiology and treatment of snakebite envenomation in west africa: case of benin. Journal of Tropical Medicine 2024(1), 8357312 (2024)

[36] Gutíerrez, J.M.: Snakebite envenoming from an ecohealth perspective. Toxicon: X 7, 100043 (2020)

[37] Silva Freitas, L., Silva, F.M., Ramires, P.F., Azevedo, G.M.G.V., Costa Batista, B.M., Anzai, R.K., Silva Pereira, I., Pissarelli, M., Andrade, F.M., Buffarini, R., et al.: Association between temperature and snakebite incidents in the state of rio grande do sul, brazil. Theoretical and Applied Climatology 156(7), 363 (2025)

[38] Silva, A.S., et al.: Predicting the drivers of bothrops snakebite incidence across brazil: A spatial analysis. Toxicon 250, 108107 (2024) 10.1016/j.toxicon.2024.108107

[39] Ochoa, C., et al.: Estimating and predicting snakebite risk in the terai region of nepal through a high-resolution geospatial and one health approach. Scientific Reports 11, 23868 (2021) 10.1038/s41598-021-03301-z

[40] Ferreira, A.A.F.e., Reis, V.P.d., Boeno, C.N., et al.: Increase in the risk of snakebites incidence due to changes in humidity levels: A time series study in four municipalities of the state of rond^onia. Revista da Sociedade Brasileira de Medicina Tropical 53, 20190377 (2020) 10.1590/0037-8682-0377-2019

[41] Musah, Y., Ameade, E.P., Attuquayefio, D.K., Holbech, L.H.: Epidemiology, ecology and human perceptions of snakebites in a savanna community of northern ghana. PLoS neglected tropical diseases 13(8), 0007221 (2019)

[42] Madsen, T., Shine, R.: Rain, fish and snakes: Climatically driven population dynamics of arafura filesnakes in tropical australia. Oecologia 124(2), 208–215 (2000) 10.1007/s004420050008

[43] Ceesay, B., Taal, A., Kalisa, M., Odikro, M.A., Agbope, D., Kenu, E.: Analysis of snakebite data in volta and oti regions, ghana, 2019. Pan African Medical Journal **40**(1) (2021)

[44] Pettorelli, N.: The Normalized Difference Vegetation Index. Oxford University Press, ??? (2013)

[45] Aglanu, L.M., Amuasi, J.H., Duah, I.K., Agbogbatey, M.K., Steinhorst, J., Adobasom-Anane, A.G., Bukari, Z., Azabu, T.J., Kreuels, B., Lalloo, D.G., et al.: Clinical presentation and management of snakebite envenoming in northern ghana. PLOS Neglected Tropical Diseases 19(12), 0013820 (2025)

[46] Molesworth, A.M., Harrison, R., David, R., Theakston, G., Lalloo, D.G.: Geographic information system mapping of snakebite incidence in northern ghana and nigeria using environmental indicators: a preliminary study. Transactions of the Royal Society of Tropical Medicine and Hygiene 97(2), 188–192 (2003)

[47] Thanappan, S., Reena, M., Saab, M., Aneley, A.D.: Identifying snake species and managing snakebite risks in construction sites. Journal of Construction and Building Materials Engineering 11(3), 32–42 (2025)

[48] Kayode, G.A., Amoakoh-Coleman, M., Brown-Davies, C., Grobbee, D.E., Agyepong, I.A., Ansah, E., Klipstein-Grobusch, K.: Quantifying the validity of routine neonatal healthcare data in the greater accra region, ghana. PLoS One 9(8), 104053 (2014)

[49] Lawson, A.B.: Bayesian Disease Mapping: Hierarchical Modeling in Spatial Epidemiology. Chapman and Hall/CRC, ??? (2018)

[50] Moraga, P.: Geospatial Health Data: Modeling and Visualization with R-INLA and Shiny. Chapman and Hall/CRC, ??? (2019)

[51] Habib, A., Abubakar, S., Abubakar, I., Larnyang, S., Durfa, N., Nasidi, A., Yusuf, P., Garnvwa, J., Theakston, R.D.G., Salako, L., et al.: Envenoming after carpet viper (echis ocellatus) bite during pregnancy: timely use of effective antivenom improves maternal and foetal outcomes. Tropical Medicine & International Health 13(9), 1172–1175 (2008)

[52] Isbister, G.K.: The critical time period for administering antivenom: golden hours and missed opportunities. Clinical Toxicology 62(5), 277–279 (2024)

[53] World Health Organization: Snakebite Envenoming: A Strategy for Prevention and Control. World Health Organization, Geneva (2019)

[54] Habib, A.G., Brown, N.I.: The snakebite problem and antivenom crisis from a health-economic perspective. Toxicon 150, 115–123 (2018)

[55] Nduwayezu, R., Kinney, H., Amuguni, J.H., Schurer, J.M.: Snakebite envenomation in rwanda: patient demographics, medical care, and antivenom availability in the formal healthcare sector. The American Journal of Tropical Medicine and Hygiene 104(1), 316 (2020)

[56] Iliyasu, G., Tiamiyu, A.B., Daiyab, F.M., Tambuwal, S.H., Habib, Z.G., Habib, A.G.: Effect of distance and delay in access to care on outcome of snakebite in rural north-eastern nigeria. Rural and remote health 15(4), 76–81 (2015)

[57] Potet, J., Beran, D., Ray, N., Alcoba, G., Habib, A.G., Iliyasu, G., Waldmann, B., Ralph, R., Faiz, M.A., Monteiro, W.M., et al.: Access to antivenoms in the developing world: a multidisciplinary analysis. Toxicon: X 12, 100086 (2021)

